# Safety of bivalent omicron-containing mRNA-booster vaccines: a nationwide cohort study

**DOI:** 10.1101/2023.01.21.23284855

**Authors:** Niklas Worm Andersson, Emilia Myrup Thiesson, Jørgen Vinsløv Hansen, Anders Hviid

## Abstract

**Background:** Safety data to support bivalent omicron-containing mRNA-booster vaccination are lacking.

**Methods:** In a Danish nationwide cohort study from 1 January 2021 to 10 December 2022, we examined the association between bivalent omicron-containing mRNA-booster vaccination as a fourth Covid-19 vaccine dose and risk of adverse events in individuals aged ≥50 years. Using incidence rate ratios estimated with Poisson regression, we compared the rates of hospital visits for 27 different adverse events in a 28-day main risk period following vaccination with a bivalent omicron-containing mRNA-booster vaccine as a fourth dose to reference period rates from day 29 after the third or fourth vaccine dose and onward. Secondary analyses included stratifying by sex, age, and vaccine type and assessing the associations using self-controlled case series and observed vs. expected cohort analyses.

**Results:** 1,740,417 individuals (mean age 67.8 years, standard deviation 10.7) received a bivalent omicron-containing mRNA-booster vaccine as a fourth dose. Fourth dose vaccination with a bivalent omicron-containing booster did not statistically significantly increase the rate of any of the 27 adverse outcomes within 28 days, nor when analyzed according to age, sex, vaccine type, or using alternative analytical approaches. However, post-hoc analysis detected signals for myocarditis (statistically significantly so in females), although the outcome was very rare and findings were based on few cases. No risk of cerebrovascular infarction was found.

**Conclusions:** Bivalent omicron-containing mRNA-booster vaccination as a fourth dose was not associated with an increased risk of 27 different adverse events in 50+-year-olds.

## INTRODUCTION

Bivalent mRNA-booster doses with either a Pfizer/BioNTech (original/omicron BA.4-5 or BA.1, *Comirnaty*) or a Moderna (original/omicron BA.1, *Spikevax*) vaccine that targets the original (ancestral) strain of SARS-CoV-2 and omicron subvariant BA.4-5 or BA.1 were authorized for use in early autumn 2022.^1–5^ These bivalent omicron-containing mRNA-booster vaccines have subsequently been recommended as single boosters after the completion of a primary vaccination series, with or without a (original) monovalent first booster (i.e., a third vaccine dose).^1–5^

In Denmark, rollout of the bivalent omicron-containing mRNA boosters started on 15 September 2022 and were recommended and offered to all individuals aged ≥50 years as well as to individuals considered at high risk of severe Covid-19. As the vaccine coverage of the third dose (i.e., first booster) has been high in Denmark (>90% for the targeted populations),^6^ the bivalent omicron-containing mRNA boosters have predominantly been administered as a fourth dose (i.e., second booster) to the general population aged ≥50 years.

Regulatory safety evaluations of the adapted bivalent mRNA-booster vaccines have primarily been based on available preauthorization clinical trials, post-marketing safety data of the monovalent mRNA vaccines, and clinical data on immunogenicity and reactogenicity of the bivalent mRNA vaccines.^5,7–14^ One recent report from the Centers of Disease Control and Prevention (CDC) of the early reported safety findings (mostly related to reactogenicity) through v-safe and the Vaccine Adverse Event Reporting System (VAERS) for the bivalent BA.4-5-containing boosters (i.e., as a ≥3 dose) appeared to be similar to those previously observed for monovalent vaccine boosters (not statistically analyzed).^15^ Consequently, data to adequately inform on the potential risk of adverse events associated with the bivalent mRNA vaccines are warranted.

In a nationwide cohort study, we investigated the association between the risk of 27 adverse events following vaccination with bivalent omicron-containing mRNA-booster vaccines as a fourth dose.

## METHODS

### Setting and study population

By use of the unique identifier assigned to all residents of Denmark upon birth or immigration, we linked data on Covid-19 vaccination, physician-assigned diagnoses, and other covariates from the nationwide healthcare registers, to construct a population-based cohort representative of the general Danish population potentially eligible for bivalent omicron-containing fourth dose booster vaccination during autumn 2022.^16–19^ As such, to be included in our cohort, an individual had to be born in 1972 or earlier (i.e. turning 50 years or older in 2022; age was defined as by 2022 minus birthyear), be a Danish resident at baseline 1 January 2021, and had to have received three prior vaccine doses with the monovalent BNT and/or MOD vaccines (or potentially AZD1222 vaccine (AstraZeneca) as part of primary vaccination)(see Table S1 in the Supplementary Appendix for details). We excluded individuals if they received the third dose within 90 days of the second dose (to ensure that the administered third dose was truly a first booster) or before 9 September 2021 (being the national rollout date of the first booster). We similarly censored fourth dose vaccinated if the vaccine was received before 15 September 2022 or within 90 days of the third dose. Vaccination status was classified in a time-varying manner during the period from 1 January 2021 to 10 December 2022. The main risk period of interest was within 28 days following the administration of the bivalent omicron-containing booster as a fourth dose while the reference period was from day 29 and after a third or fourth dose and onward (see Figure S1 for a graphical illustration of the study design).

According to Danish law, register-based research is exempt from ethics committee approval.

### Outcomes

We included 27 different adverse events, adapted from prioritized lists of adverse events of special interest for the Covid-19 vaccines.^20–22^ During follow-up, we identified outcome events as any hospital visits where an outcome diagnosis was recorded as the primary reason for seeking hospital care (see Table S2 for specific International Classification of Diseases, 10th revision-codes).^16^ We included only incident events by excluding individuals who had a history of the respective studied outcome during a washout period from 1 January 2018 to day 28 after the third dose. The date of admission served as the event date. Each of the 27 outcomes was studied separately.

### Statistical analysis

Individuals were followed from day 29 after the third dose until first outcome event, emigration/disappearance, death, receipt of a fifth vaccine dose, or end of the study period. Individuals could contribute with person-time during both the main 28-day risk and the two reference periods; number of events and person-time from the two reference periods were aggregated. We used Poisson regression to estimate adjusted incidence rate ratios (IRRs), with corresponding 95% confidence intervals (CI), comparing the outcome rates during the risk period to the reference period rates. The model was adjusted for sex, age groups (50-64, 65-79, and ≥80 years), ethnicity (Nordic, Western, non-Western), region of residence, considered at high risk of severe Covid-19 (yes/no), calendar time (3-month-bins), and number of comorbidities (defined as asthma, other chronic respiratory disorders, chronic cardiac disorders, renal disorders, diabetes, autoimmune-related disorders, epilepsy, malignancies, and psychiatric disorders).

In secondary analysis, we analyzed the associations by sex, age, and the individual bivalent booster vaccine type, assessed the risk of severe outcome events, as defined by hospital contacts of ≥5 hours, including both primary and secondary the outcome definition, and altered the main risk period to 7, 14, and 90 days since fourth dose vaccination. Further, the robustness of the results from the primary analysis were evaluated by use of two alternative analytical approaches. First, nested within our study cohort, we conducted a self-controlled case series (SCCS) analysis;^23^ in this design, only individuals experiencing an (incident) event were included, and we used conditional Poisson regression to estimate IRRs according to vaccination status. For the SCCS analysis, we excluded the 14 days prior to fourth dose vaccination from the reference period (to account for potential event-dependent exposure bias). An advantage of the SCCS design is that time-periods are compared within study individuals which provides indirect adjustment of confounders not varying over the study period such as many lifestyle- and socioeconomic factor or comorbidities. Second, we compared the ratio between the observed rate of the studied outcome with the expected rate with 95% CIs obtained from the Poisson distribution. The expected rates were estimated using indirect age- and sex standardization of the historic background rates within the Danish general population from 1 January 2015 to 31 December 2019, emigration/disappearance.

In post-hoc analysis, we specifically addressed the risk of cerebrovascular infarction, myocarditis, and pericarditis separately, including in sex and age subgroups as well as for the individual bivalent booster vaccine types. We used SAS version 9.4 and R version 4.0.2 for data management, while statistical analysis was accomplished using R version 4.0.2.

## RESULTS

### Study population

The study cohort comprised a total of 2,225,567 individuals (mean age 66.9 years, standard deviation [SD] 11.0 years and 52.0% were females) who had received at least three Covid-19 vaccine doses (91.2% received a monovalent BNT vaccine as the third dose), of whom 1,740,417 (78.2%, mean age 68.7 years, SD 10.7 years) received a fourth dose with a bivalent omicron-containing mRNA-booster vaccine during 15 September 2022 to 10 December 2022 (Table 1). The bivalent BNT BA.4-5-containing booster was the most commonly administered (1,157,754 vaccine recipients, 66.5%), followed by the BNT and MOD BA.1-containing vaccines (526,397, 30.2% and 56,266, 3.3%, respectively). Except for chronic cardiac disorders (almost 10.0%), most comorbidities had a prevalence of ≤5.0% within the cohort.

**Table 1.**
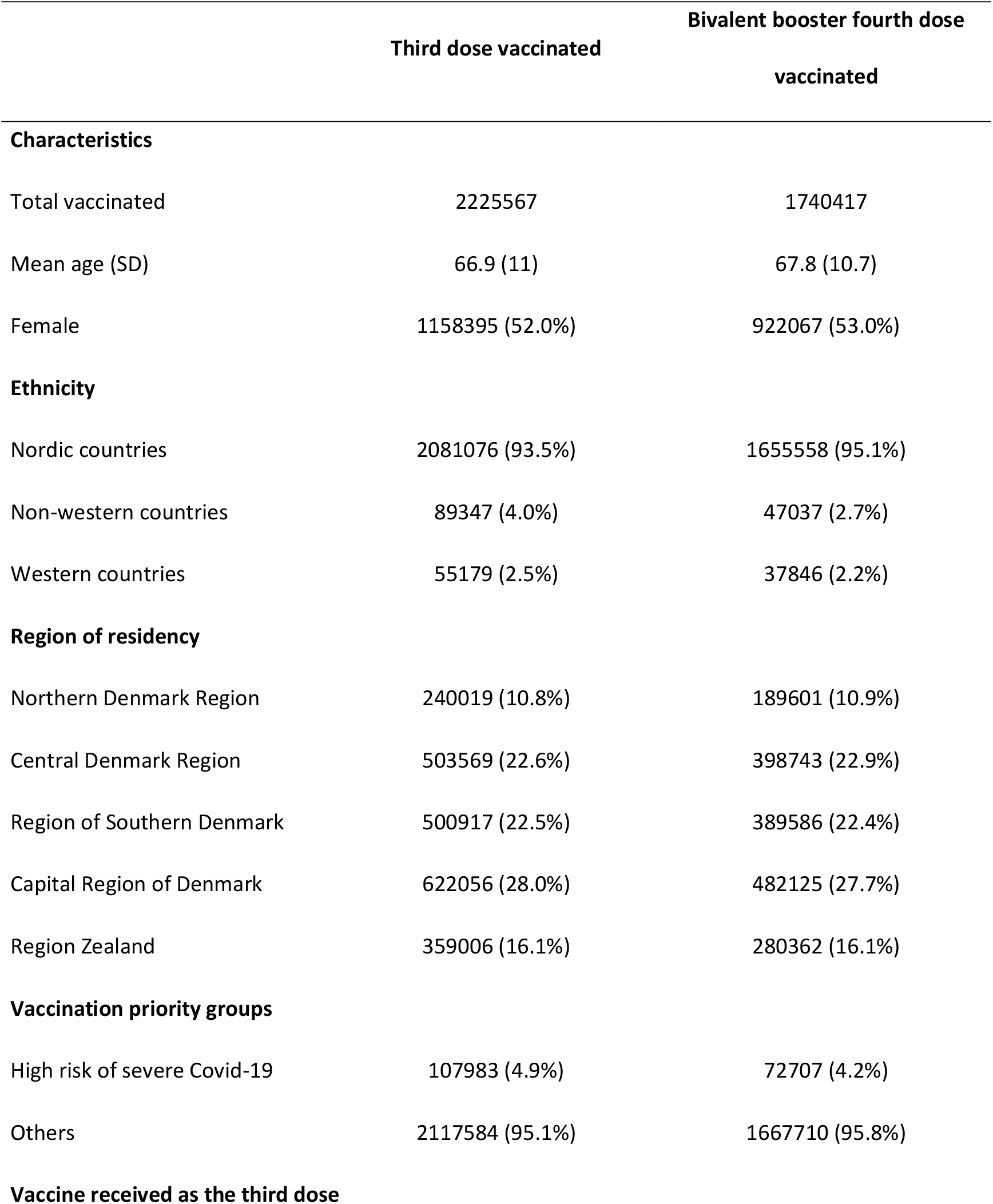
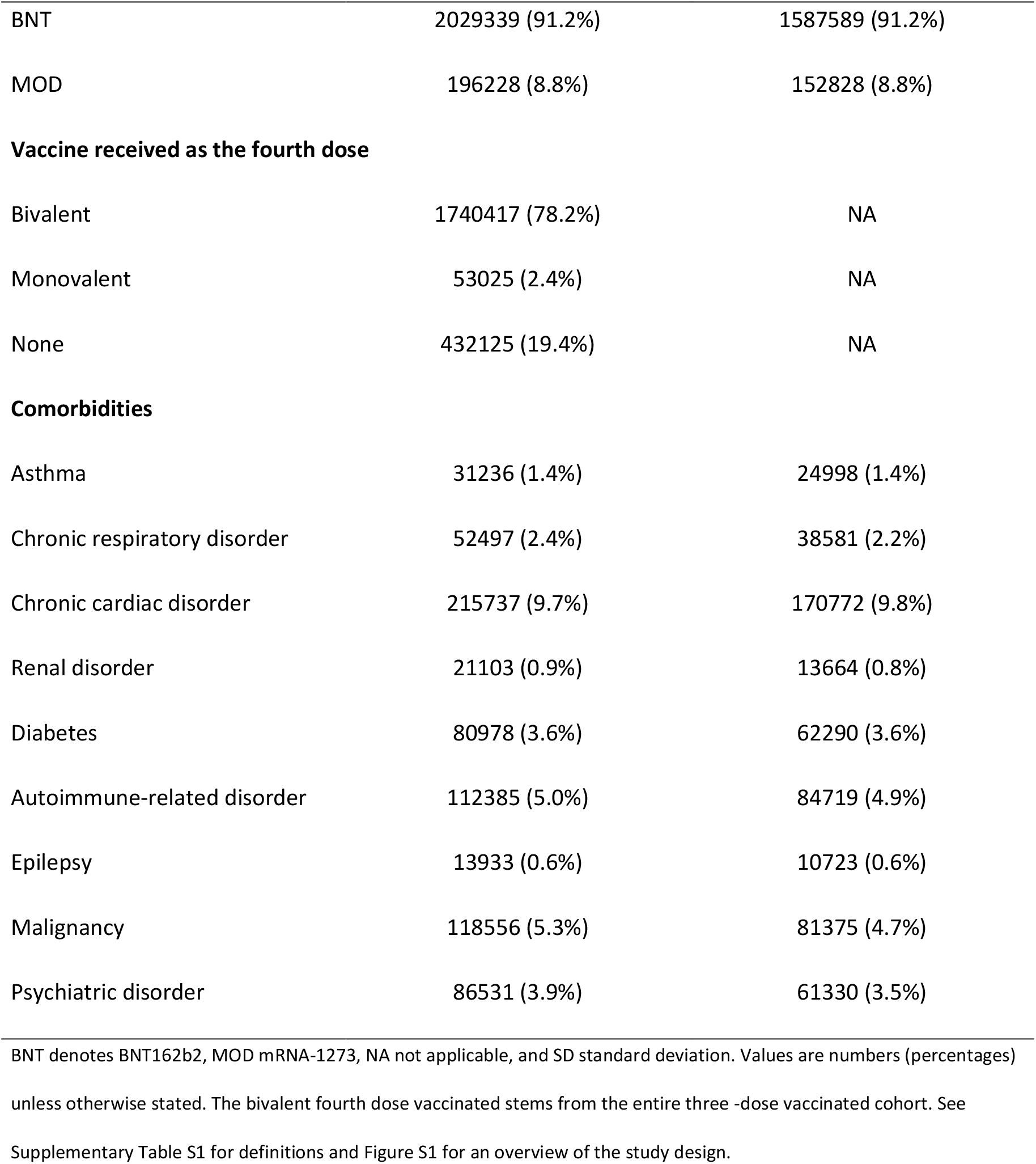
Characteristics of all individuals vaccinated with a monovalent mRNA vaccine as a third dose and a bivalent omicron-containing vaccine as a fourth dose in Denmark.

### Risk of adverse events

Fourth dose vaccination with a bivalent omicron-containing mRNA-booster vaccine was not associated with a statistically significant increased rate of hospital visits for any of the 27 different adverse events within 28 days after vaccination as compared with the reference period rates (Figure 1). Similarly, no significant associations were found when stratifying by sex and age groups (Table 2) nor by the type of bivalent booster received (Figure 2 and Table S3). Using the alternative analytical approaches of SCCS (Table S4) and observed vs. expected (Table S5) analyses showed comparable results to our main analysis (except for a rate ratio of 2.22, 95% CI 1.27 to 3.60 for subacute thyroiditis based on 16 observed vs. 7.2 expected cases). Comparing the risk of severe adverse events, defined as hospital visits of ≥5 hours, showed close to similar results of the main analysis (Table S6; no cases of subacute thyroiditis requiring hospitalization for ≥5 hours were observed during the 28-day period following fourth dose vaccination). Including both primary and secondary diagnoses in the outcome definitions did not change the results (Table S7). Our secondary analysis with use of different risk periods of 7, 14, and 90 days following fourth dose vaccination showed an increased risk of myocarditis or pericarditis (IRR 1.88, 95% CI 1.13 to 3.11) within 14 days only (Table S8); myocarditis and pericarditis were examined separately in post-hoc analyses.

**Table 2.**
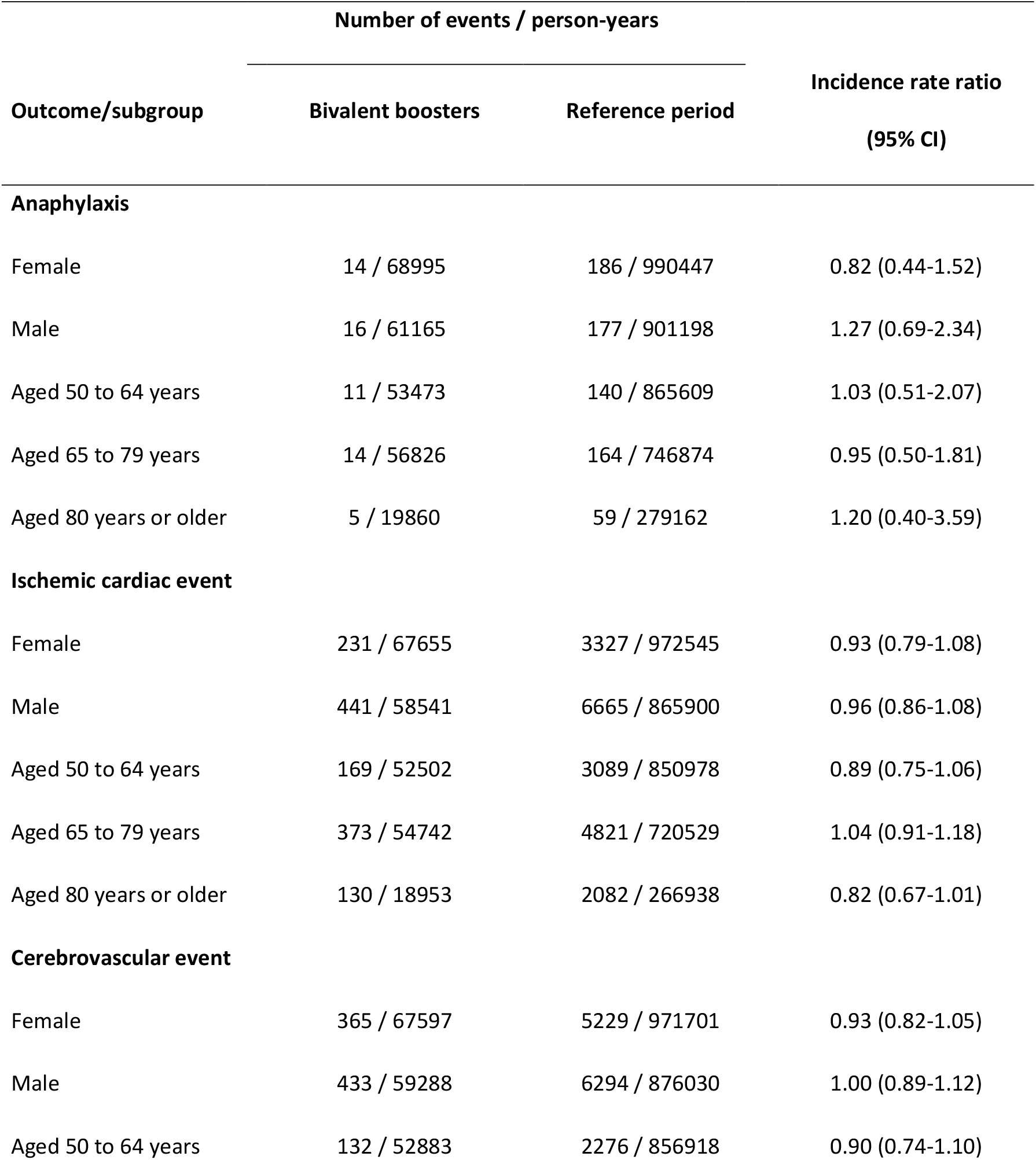
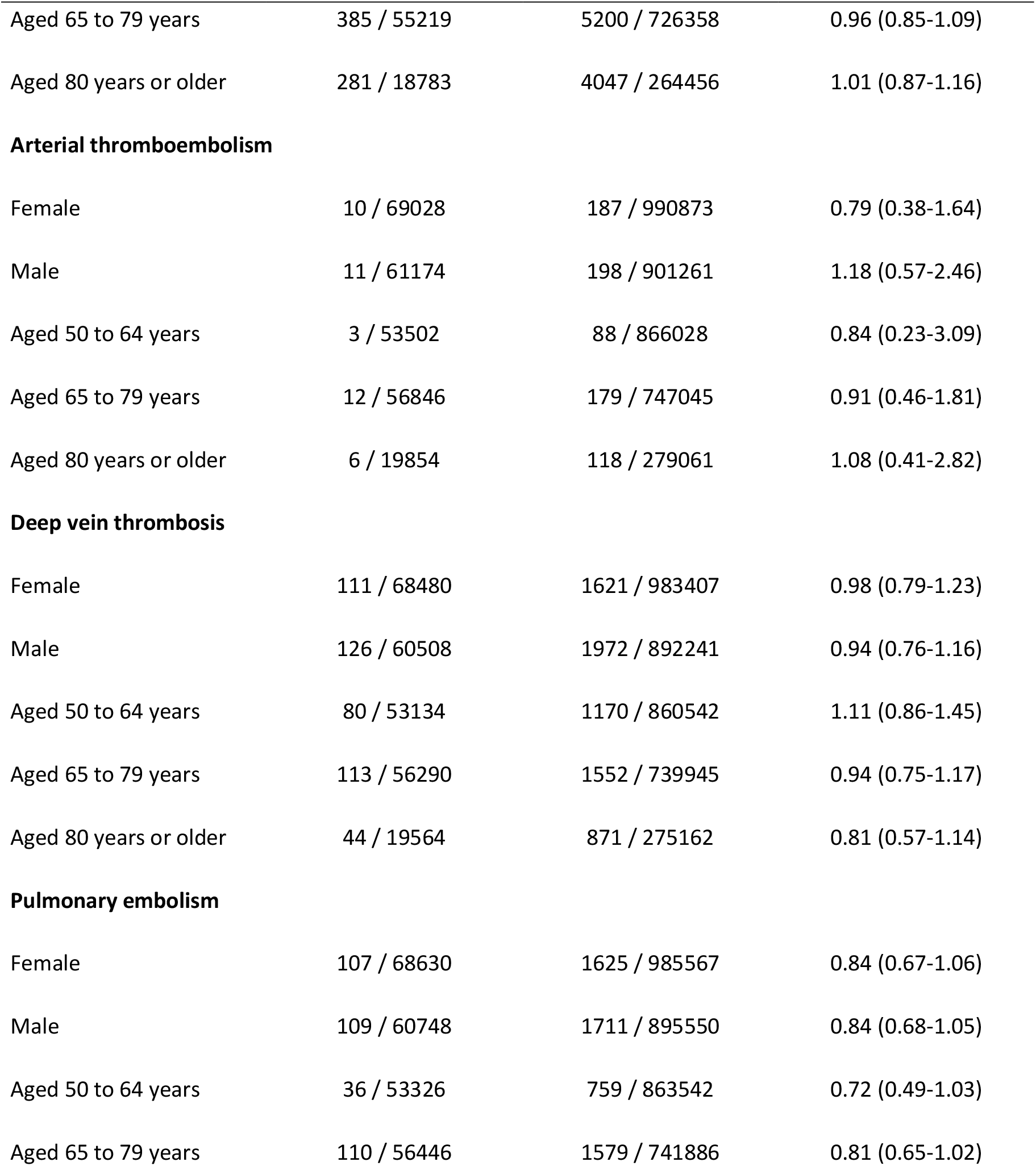
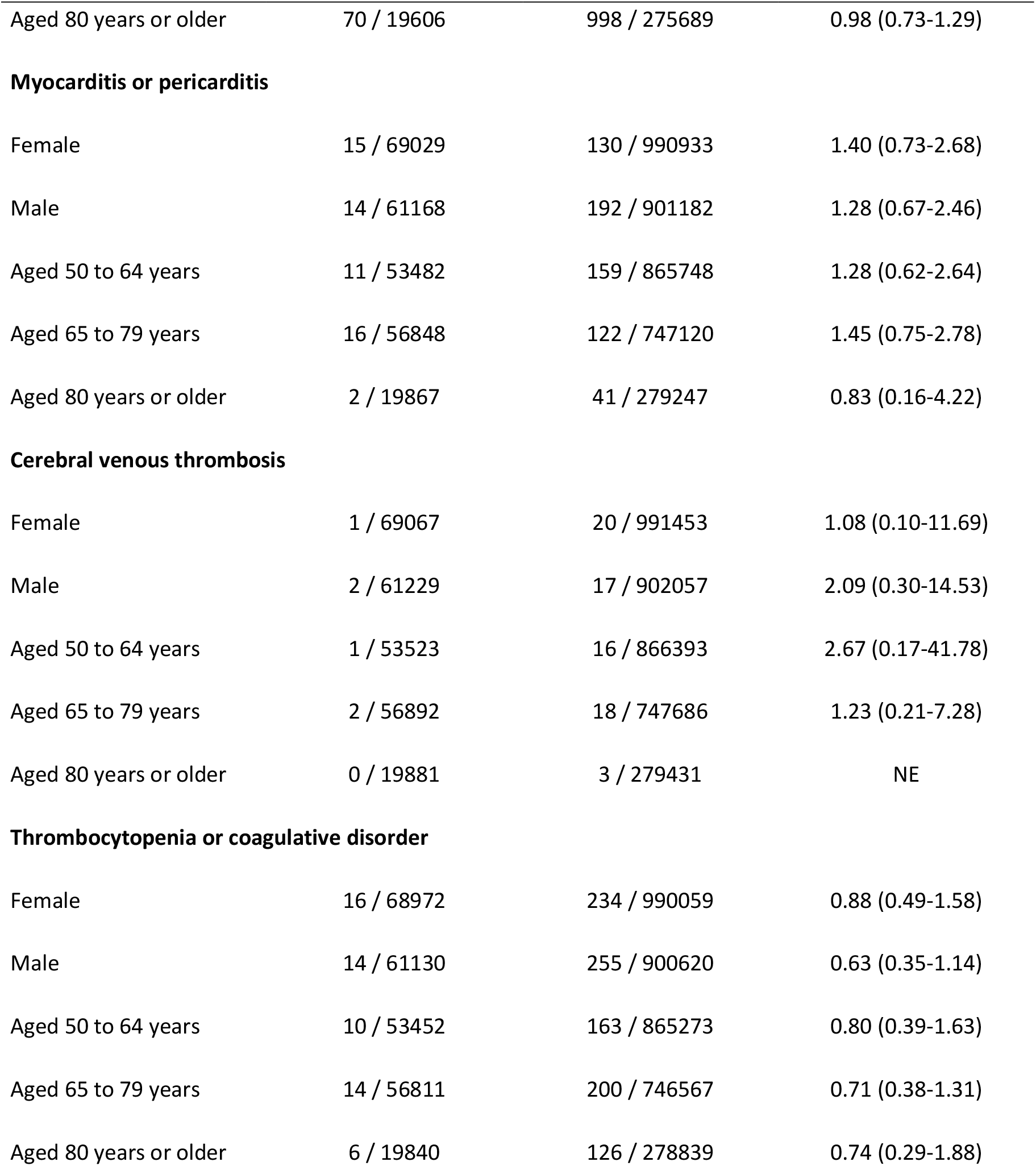
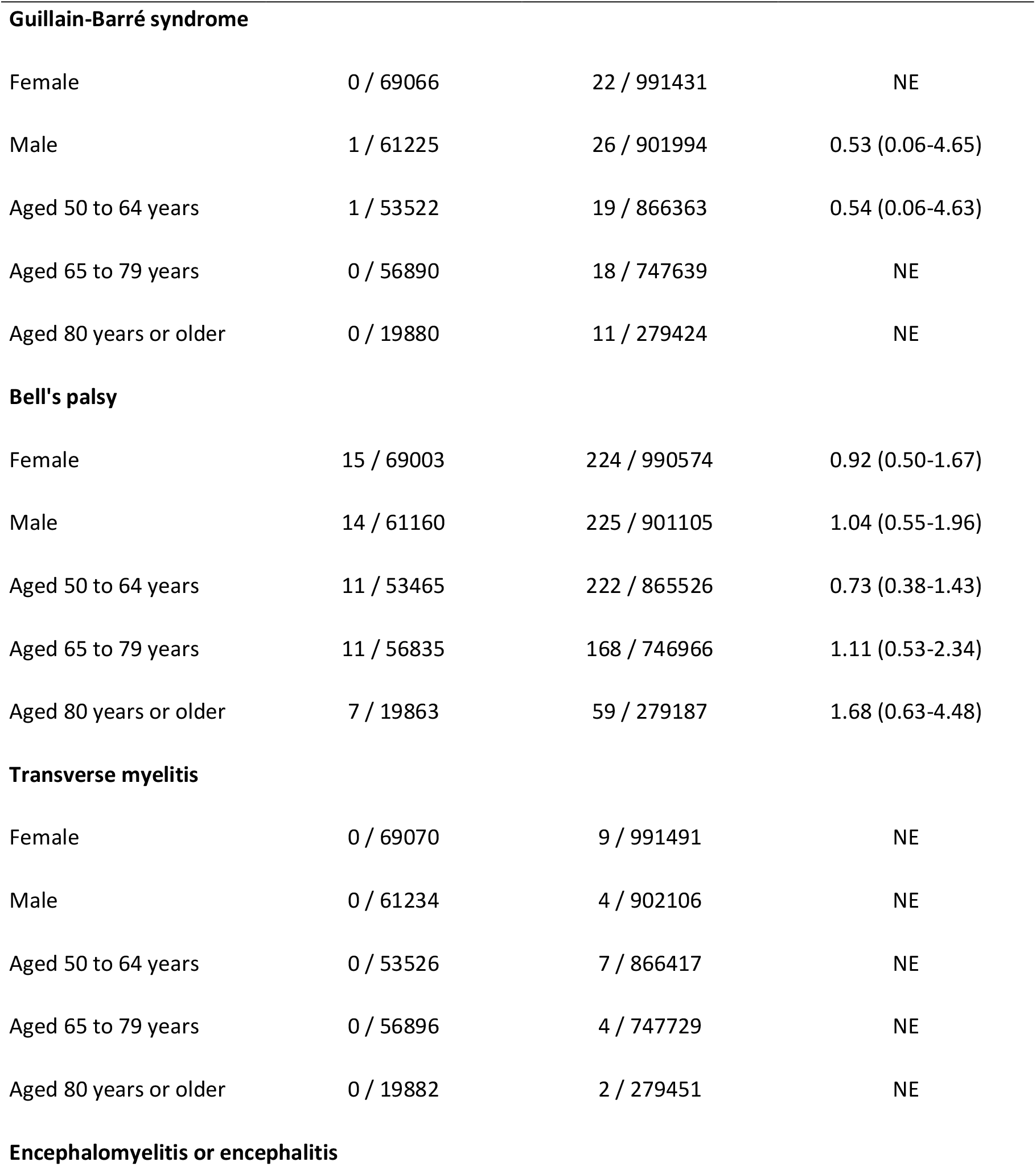
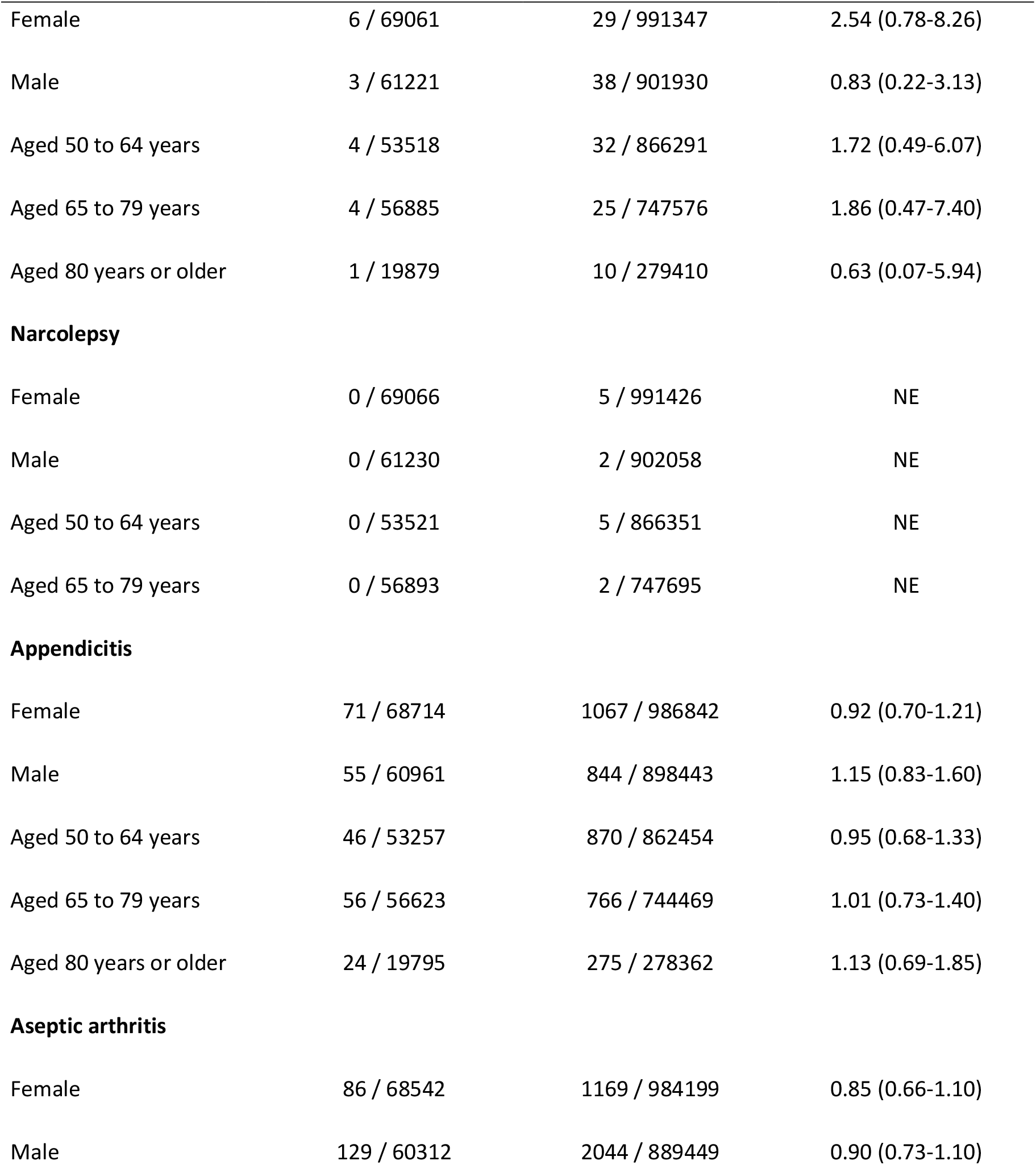
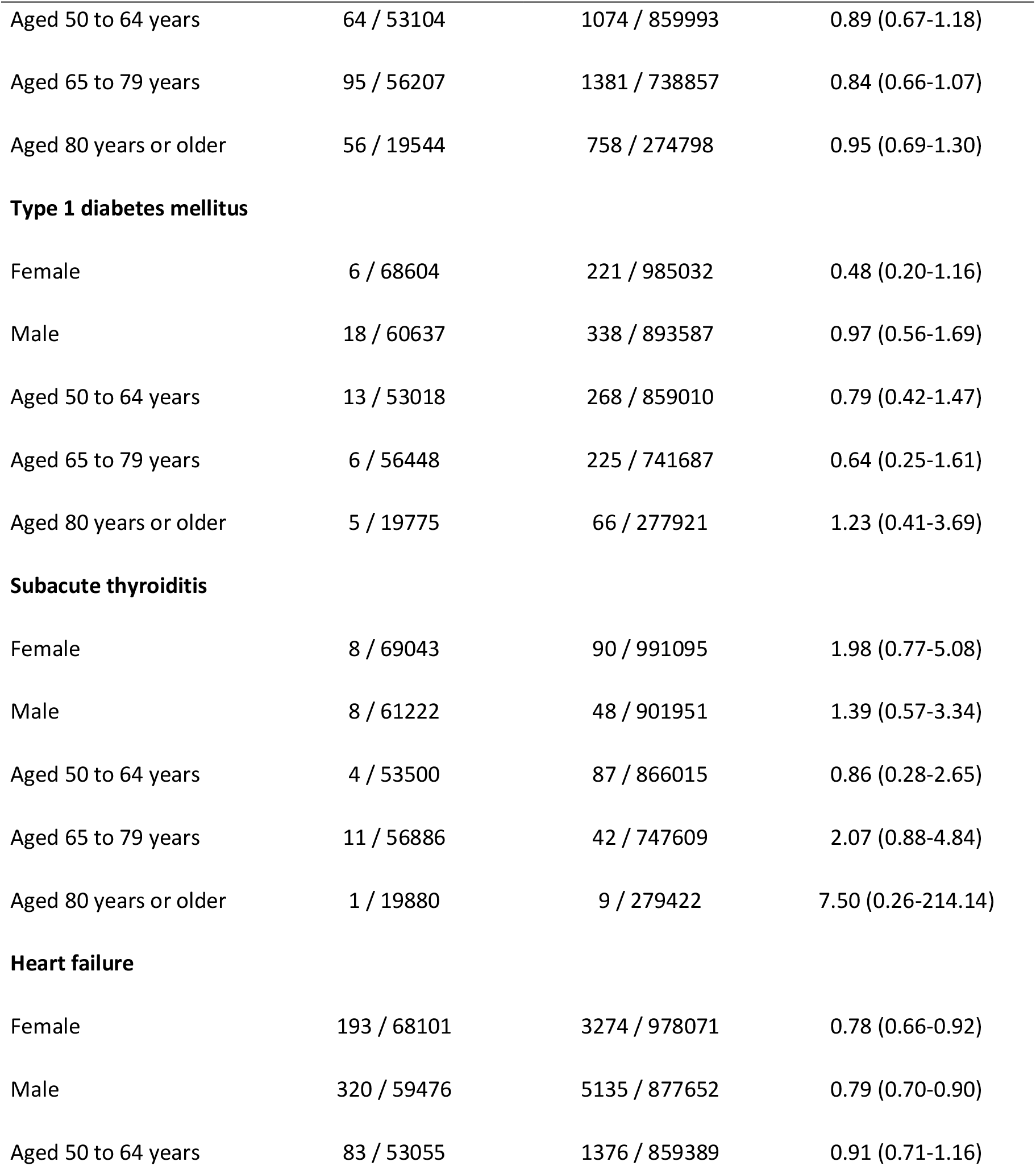
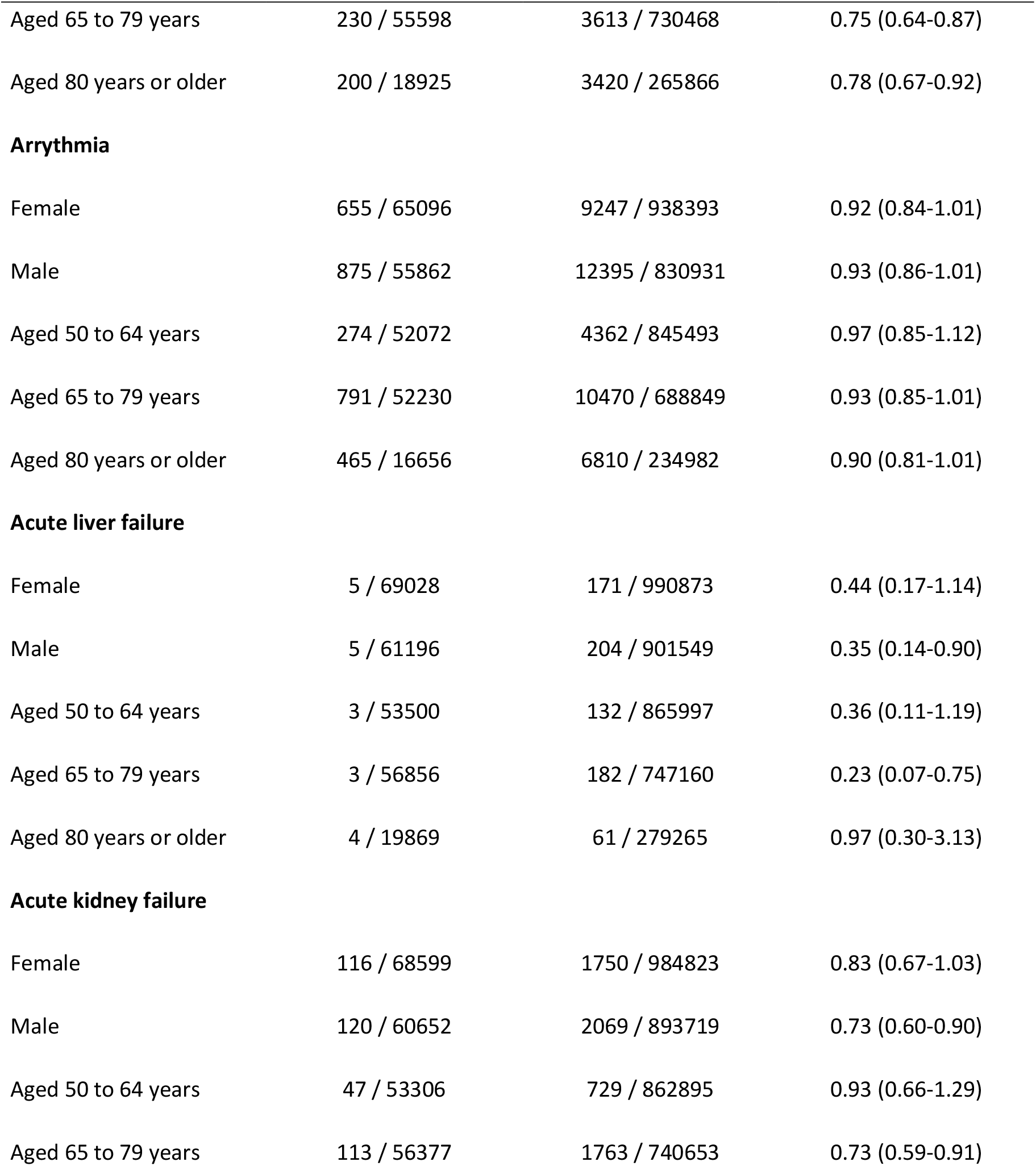
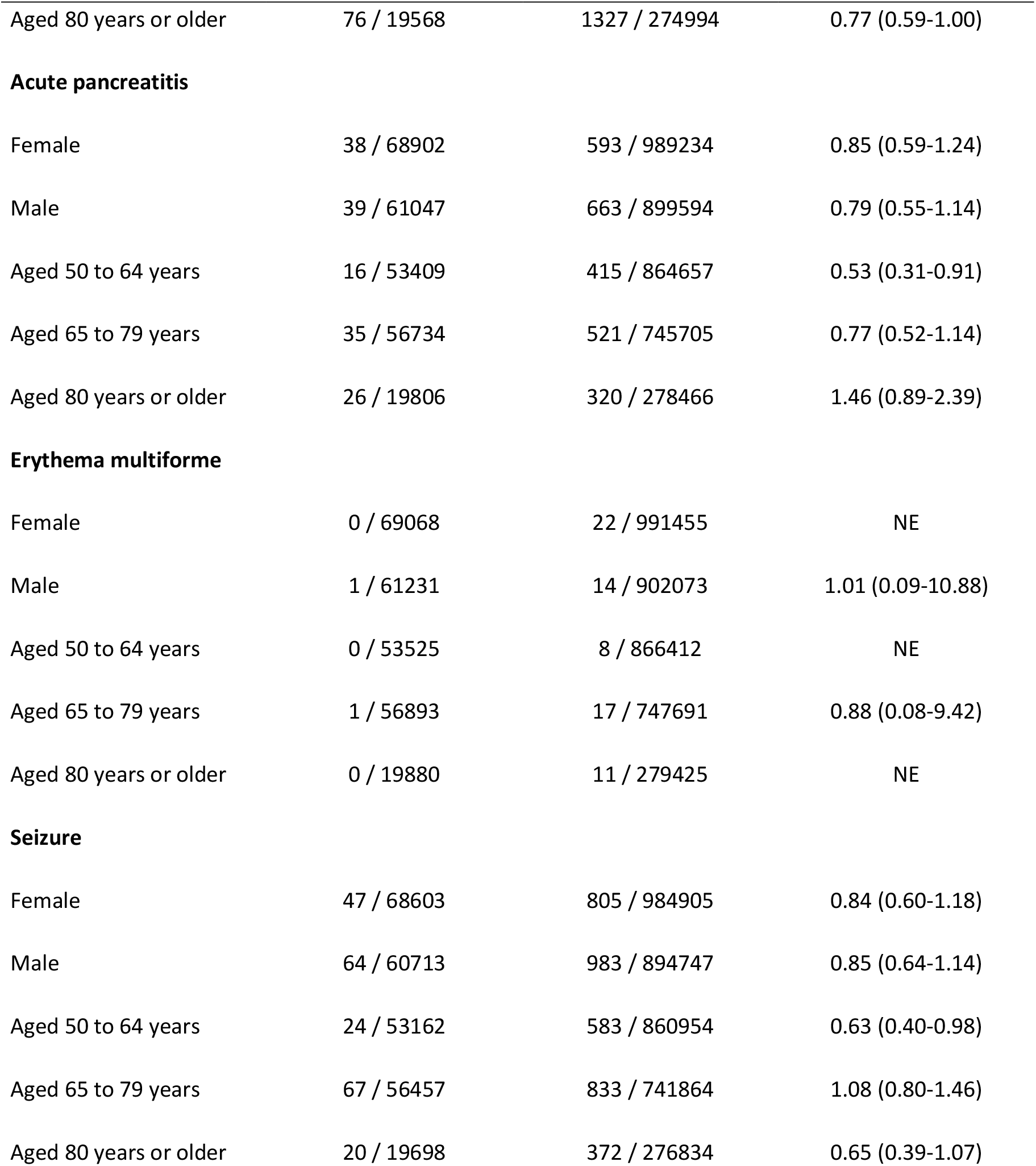
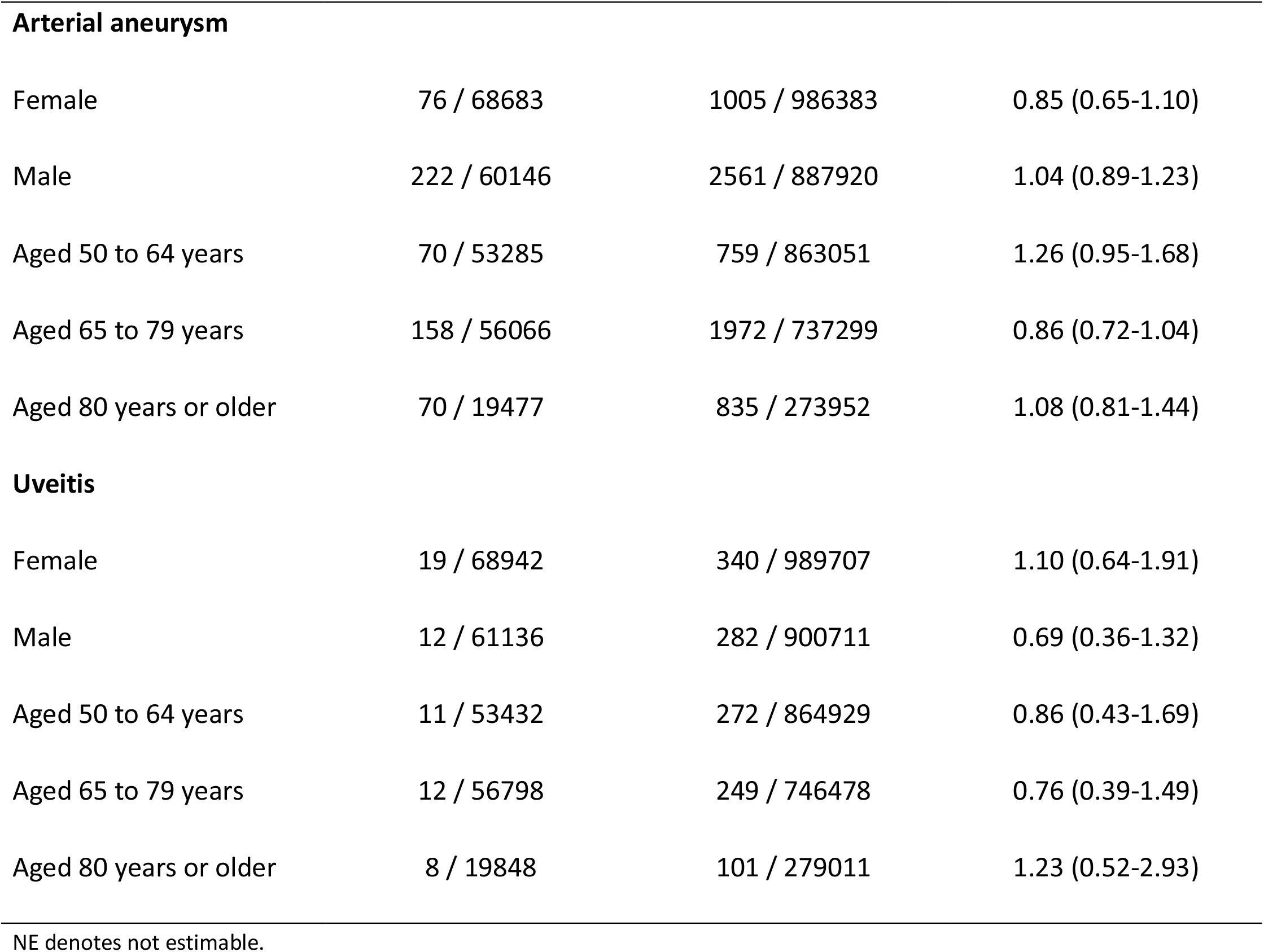
Risk of adverse events within 28 days after vaccination with a bivalent omicron-containing mRNA-booster vaccine as the fourth dose in Danish 50+-year-olds during 15 September 2022 to 10 December 2022 by sex and age.

**Figure 1.**
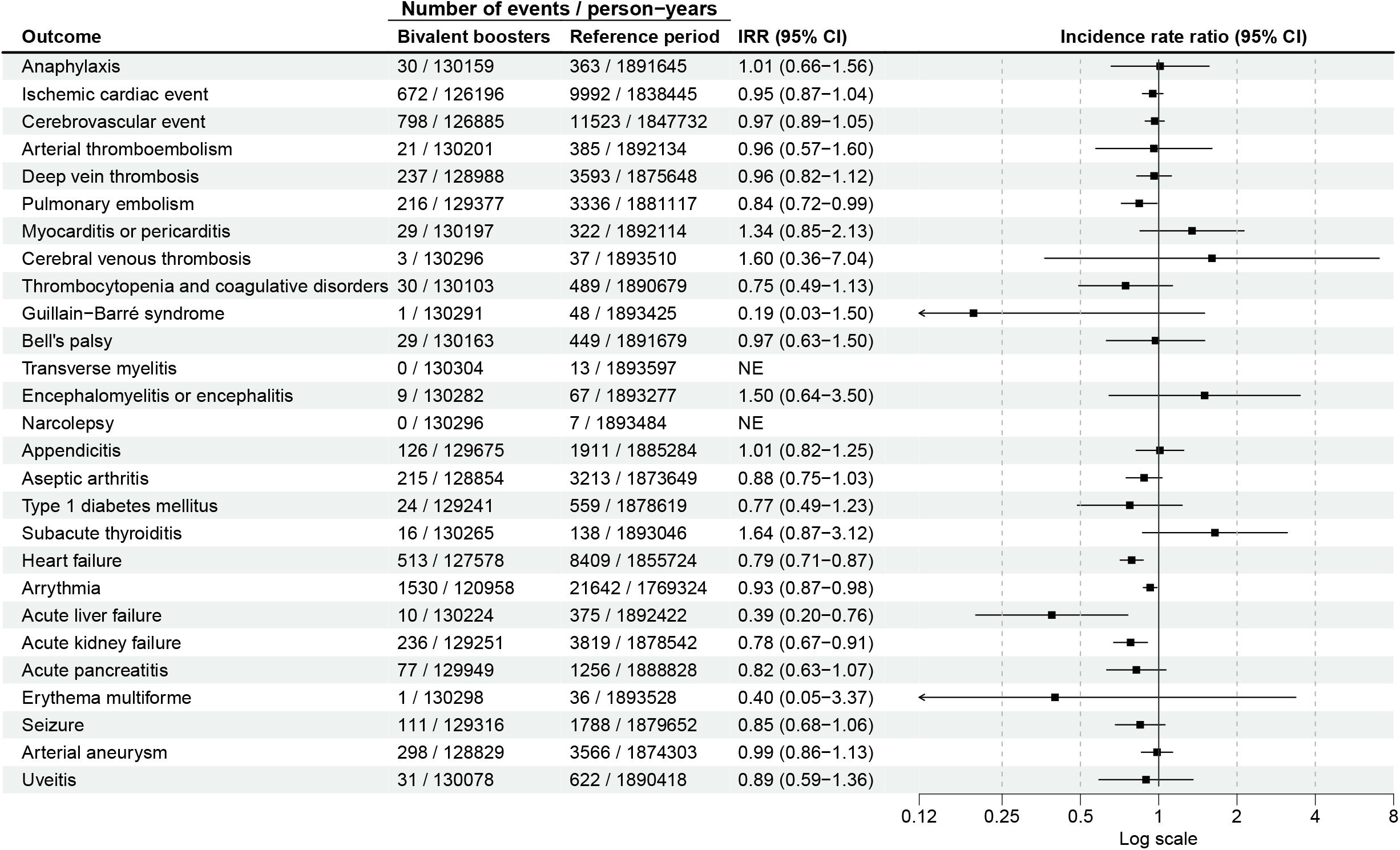
Risk of adverse events within 28 days after vaccination with a bivalent omicron-containing mRNA-booster vaccine as the fourth dose in Danish 50+-year-olds during 15 September 2022 to 10 December 2022. NE denotes not estimable.

**Figure 2.**
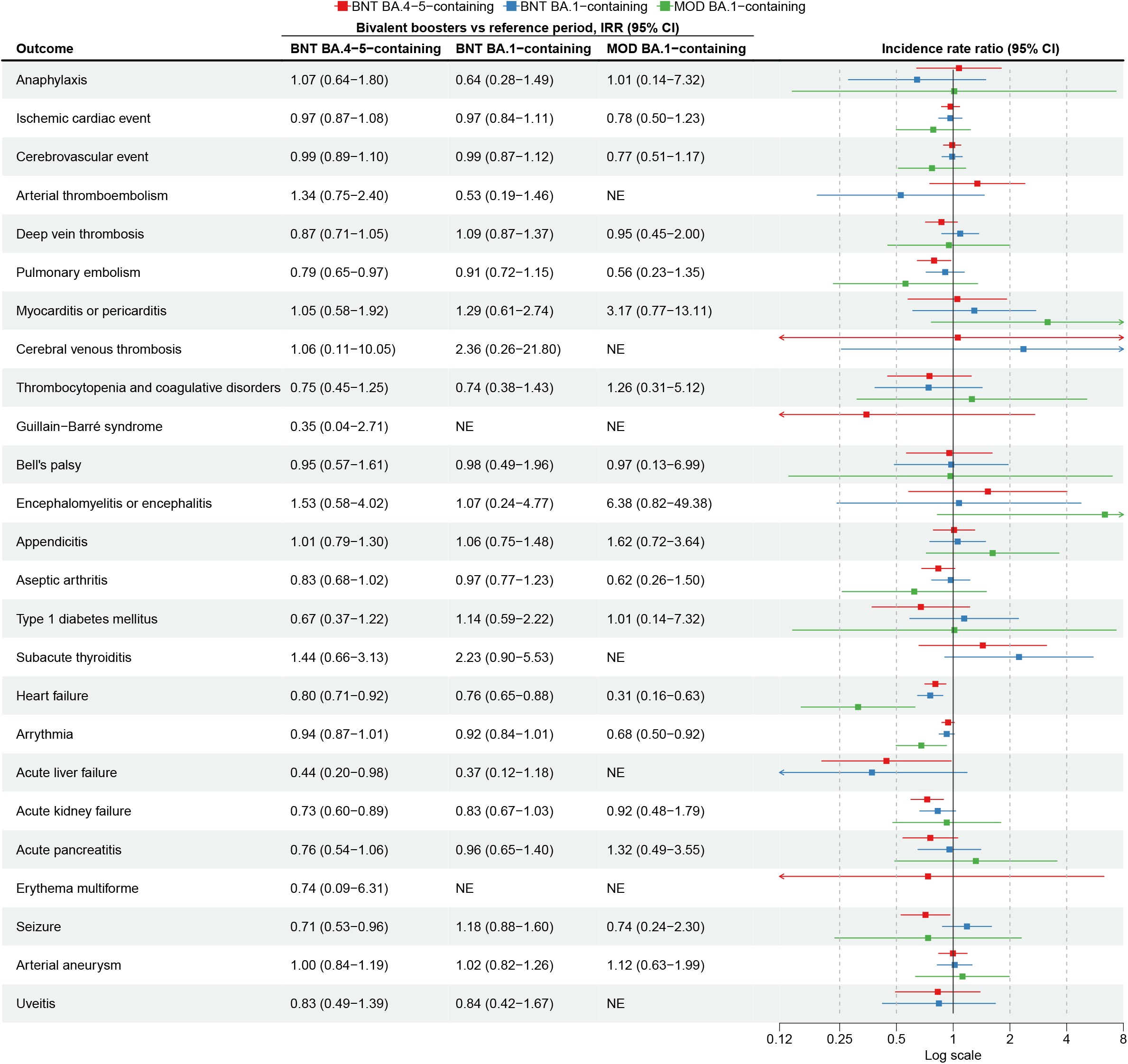
Risk of adverse events within 28 days after vaccination with a bivalent omicron-containing mRNA-booster vaccine as the fourth dose in Danish 50+-year-olds during 15 September 2022 to 10 December 2022 by type of vaccine received. ^**a**^BNT denotes BNT162b2, MOD mRNA-1273, and NE not estimable. ^a^The corresponding number of events and person-years are presented in Supplementary Table S3.

### Post-hoc analysis

No association between risk of cerebrovascular infarction and fourth dose bivalent booster vaccination was observed (IRR 0.95, 95% CI 0.87 to 1.05), including by sex, age, and type of bivalent booster (Table 3 and Table S9). We observed a total of 9 cases of myocarditis and 22 cases of pericarditis within 28 days following a fourth dose with a bivalent omicron-containing booster (equal to incidence rates of 5.3 and 13.0 cases within 28 days per 1,000,000 vaccinated, respectively; reference period incidence rates were 2.6 and 10.7 per 28 days among 1,000,000 individuals, respectively). The corresponding IRR was 2.65 (95% CI 1.01 to 6.97) for myocarditis and 1.21 (0.72 to 2.03) for pericarditis. Statistically significant associations with myocarditis were observed for females and the BNT BA.4-5-containing and MOD BA.1-containing boosters; the point estimate was similarly elevated for the BNT BA.1-containing booster but not statistically significant. Examining the associated risk of myocarditis with the individual vaccine types by sex and age groups showed statistically significant associations in females for all three vaccine types and in individuals aged 50 to 64 years who had been vaccinated with the MOD BA.1-containing booster (Table S9). However, all subgroup analyses of myocarditis risk were based on ≤6 cases among fourth dose vaccinated. No associations with pericarditis were found in any analysis.

**Table 3.** Risk of cerebrovascular infarction, myocarditis, and pericarditis following fourth dose vaccination with a bivalent omicron-containing booster within 28 days in Danish 50+-year-olds during 15 September 2022 to 10 December 2022 including by sex, age, and vaccine type.

| Outcome/subgroup | Bivalent boosters |  | Reference period |  | Incidence rate ratio<br>(95% CI) |
| --- | --- | --- | --- | --- | --- |
|  | Events/<br>person-years | Rate/<br>1,000,000<sup>a</sup> | Events/ person-years | Rate/<br>1,000,000<sup>a</sup> |  |
| Cerebrovascular infarction (including TIA) |  |  |  |  |  |
| All | 644 / 127304 | 387.8 | 9687 / 1853451 | 400.7 | 0.95 (0.87-1.05) |
| Females | 290 / 67794 | 327.9 | 4317 / 974392 | 339.6 | 0.91 (0.79-1.05) |
| Males | 354 / 59511 | 456.0 | 5370 / 879059 | 468.3 | 0.99 (0.88-1.13) |
| Aged 50 to 64 years | 113 / 52983 | 163.5 | 1910 / 858398 | 170.6 | 0.96 (0.78-1.20) |
| Aged 65 to 79 years | 315 / 55419 | 435.7 | 4426 / 728948 | 465.5 | 0.94 (0.82-1.07) |
| Aged 80 years or older | 216 / 18902 | 876.0 | 3351 / 266105 | 965.4 | 0.98 (0.83-1.15) |
| BNT BA.4-5-containing | 377 / 84259 | 343.0 | 9687 / 1853451 | 400.7 | 0.97 (0.86-1.09) |
| BNT BA.1-containing | 246 / 38856 | 485.3 | 9687 / 1853451 | 400.7 | 0.98 (0.85-1.12) |
| MOD BA.1-containing | 21 / 4189 | 384.3 | 9687 / 1853451 | 400.7 | 0.83 (0.54-1.29) |
| Myocarditis |  |  |  |  |  |
| All | 9 / 130288 | 5.3 | 64 / 1893405 | 2.6 | 2.65 (1.01-6.97) |
| Females | 6 / 69066 | 6.7 | 29 / 991433 | 2.2 | 3.89 (1.05-14.50) |
| Males | 3 / 61223 | 3.8 | 35 / 901972 | 3.0 | 1.60 (0.36-7.07) |
| Aged 50 to 64 years | 4 / 53520 | 5.7 | 30 / 866339 | 2.7 | 3.29 (0.75-14.46) |
|  | Events/<br>person-years | Rate/<br>1,000,000<sup>a</sup> | Events/ person-years | Rate/<br>1,000,000<sup>a</sup> |  |
| Aged 65 to 79 years | 4 / 56889 | 5.4 | 27 / 747644 | 2.8 | 1.79 (0.45-7.08) |
| Aged 80 years or older | 1 / 19880 | 3.9 | 7 / 279421 | 1.9 | 10.13 (0.23-437.19) |
| BNT BA.4-5-containing | 6 / 85859 | 5.4 | 64 / 1893405 | 2.6 | 3.74 (1.16-12.05) |
| BNT BA.1-containing | 2 / 40146 | 3.8 | 64 / 1893405 | 2.6 | 2.59 (0.52-12.94) |
| MOD BA.1-containing | 1 / 4283 | 17.9 | 64 / 1893405 | 2.6 | 12.39 (1.46-105.47) |
| Pericarditis |  |  |  |  |  |
| All | 22 / 130215 | 13.0 | 265 / 1892351 | 10.7 | 1.21 (0.72-2.03) |
| Females | 11 / 69036 | 12.2 | 104 / 991018 | 8.0 | 1.20 (0.57-2.50) |
| Males | 11 / 61179 | 13.8 | 161 / 901332 | 13.7 | 1.22 (0.59-2.53) |
| Aged 50 to 64 years | 8 / 53490 | 11.5 | 134 / 865857 | 11.9 | 1.09 (0.48-2.48) |
| Aged 65 to 79 years | 13 / 56856 | 17.5 | 96 / 747218 | 9.8 | 1.47 (0.71-3.05) |
| Aged 80 years or older | 1 / 19869 | 3.9 | 35 / 279276 | 9.6 | 0.43 (0.05-3.67) |
| BNT BA.4-5-containing | 14 / 85811 | 12.5 | 265 / 1892351 | 10.7 | 0.85 (0.43-1.71) |
| BNT BA.1-containing | 7 / 40123 | 13.4 | 265 / 1892351 | 10.7 | 1.12 (0.48-2.64) |
| MOD BA.1-containing | 1 / 4281 | 17.9 | 265 / 1892351 | 10.7 | 1.82 (0.25-13.31) |

## DISCUSSION

In a nationwide cohort analysis, we evaluated the rate of 27 adverse events associated with the bivalent omicron-containing mRNA-booster vaccines received as the fourth dose in 1,740,417 individuals aged ≥50 years. We found no support for an increased risk of adverse events following vaccination with a bivalent omicron-containing mRNA booster as a fourth dose.

Our post-hoc analysis did reveal an excess of hospital visits due to myocarditis in females. However, myocarditis after bivalent omicron-containing mRNA-booster vaccination was a very rare event (a total of 9 cases) during the 28-day post-vaccination period of follow-up. In absolute numbers, we observed a rate of 5.3 cases in the 28-day post-vaccination period per 1,000,000 vaccinated, compared to a reference period incidence rate of 2.6 cases in 28-days per 1,000,0000 individuals. Previous reported rates of myocarditis in adults following vaccination with the monovalent mRNA vaccines were highest in younger males.^24–26^ Five reports of myocarditis events following bivalent booster vaccination (not statistically analyzed) was received through VAERS in the recent CDC report on 22.6 million bivalent booster doses administered in the US to individuals aged ≥12 years.^15^

The CDC and the Food and Drug Administration (FDA) recently identified possible safety concerns of increased risk of cerebrovascular infarction following bivalent BNT BA.4-5-containing booster vaccination in adults aged ≥65 years due to detected preliminary safety signal by the CDC Vaccine Safety Datalink surveillance system.^27^ We found no support for this notion, including no associations in any age, sex, and vaccine type subgroup analyses. Particularly, the upper 95% CI-bound from our analyses based on 1,157,754 fourth dose BNT BA.4-5-containing booster vaccinated individuals were inconsistent with relative increased risks of cerebrovascular infarction larger than 9% (IRR 0.97, 95% CI 0.86-1.09).

The robustness of the results from our main analysis is supported by SCCS analyses taking time invariant confounding into account and comparisons to historical rates. Furthermore, the use of nationwide demography- and health registers in a setting with universal access to free healthcare reduces concern about selection- and information bias. However, a limitation of our study is that we cannot exclude differences in the ascertainment of adverse events between compared periods. The active comparative nature of our design, where we compare the 28-day period rates following fourth dose vaccination with reference periods rates from day 29 after vaccination, mitigates larger differences in outcome ascertainment as opposed to a comparison with unvaccinated period rates. However, ascertainment bias cannot be completely excluded; specifically, an increased awareness of any symptoms related to known adverse events (e.g., myocarditis) in the weeks following fourth dose vaccination relative to the reference periods with longer time since vaccination. However, this would bias the results towards increased risks in contrast to what we observe.

A strength of our study is the high statistical precision of many of the evaluated associations. For 19 of the 27 adverse events analyzed, the upper confidence interval bounds excluded 1.50 or higher, suggesting that moderate-to-large relative risk increases are unlikely.

Owing to the nationwide-based design, our findings have a high degree of generalizability to similar populations. In contrast, these results may only indirectly support safety evaluations of bivalent omicron-containing booster vaccination in other scenarios such as in individuals younger than 50 years old or other specific subgroups that were not studied.

## CONCLUSION

Bivalent omicron-containing mRNA-booster vaccines administered as a fourth dose were not associated with an increased risk of 27 different adverse events in a general population of 50+-year-olds. These results provide reassuring support for the safety of bivalent booster vaccination, and provides critical and timely insight to patients, clinicians, and regulatory authorities.

## Supporting information

Table S1 in the Supplementary Appendix

## Data Availability

No additional data available. Owing to data privacy regulations in Denmark, the raw data cannot be shared.

## Contributors

All authors conceptualized the study, interpreted the results and critically reviewed the manuscript; NA drafted the manuscript; NA, ET, and JH carried out the statistical analyses. AH supervised the study. NA had full access to all the data in the study and take responsibility for the integrity of the data and the accuracy of the data analyses.

## Transparency

The lead author (the manuscript’s guarantor) affirms that this manuscript is an honest, accurate, and transparent account of the study being reported; that no important aspects of the study have been omitted; and that any discrepancies from the study as planned (and, if relevant, registered) have been explained.

## Ethics

The analyses were performed as surveillance activities analyses as part of the advisory tasks of the governmental institution Statens Serum Institut (SSI) for the Danish Ministry of Health. SSI’s purpose is to monitor and fight the spread of disease in accordance with section 222 of the Danish Health Act. According to Danish law, national surveillance activities conducted by SSI do not require approval from an ethics committee. Both the Danish Governmental law firm and the compliance department of SSI have approved that the study is fully compliant with all legal, ethical, and IT-security requirements and there are no further approval procedures required for such studies.

## Dissemination to participants and related patient and public communities

Studied participants were anonymised in the utilised data sources; and therefore, direct dissemination to study participants is not possible. The study results will be disseminated to the public and health professionals by a press release written using layman’s terms.

