## Supplementary material for "Safety of bivalent omicron-containing mRNA-booster vaccines: a nationwide cohort study": Table S1 in the Supplementary Appendix

### Supplementary Table S1. Eligibility criteria and covariates definition.

| Variable | Details |
|---|---|
| **Eligibility criteria** | |
| Age of 50 years or older | *The Civil Registration System*[1]. Defined by time birthyear and year 2022. National vaccine policy of fourth dose rollout, individuals had to be born in year 1972 or earlier. |
| Vaccinated with a primary course and one booster Covid-19. | *The Danish Vaccination Register*[2]. Defined as registered received three previous Covid-19 vaccines with the BNT162b2 and/or mRNA-1273 as well as and/or AZD1222 vaccine (the latter as part of the primary vaccination course only). Other vaccine types were rare within our study population. During the national rollout of the primary vaccination course, vaccination with the AZD1222 vaccine was halted in Denmark in early March 2021. |
| Third vaccine dose not received earlier than 9 September 2021[a] | *The Danish Vaccination Register*[2]. First day of national rollout of the third (i.e., first booster vaccine dose) to the general Danish population. |
| Third vaccine dose not received within 90 days of the second dose[b] | *The Danish Vaccination Register*[2]. As vaccination with a third vaccine dose within 90 days of the primary vaccination schedule does not constitute as a booster dose per se, but rather a part of the primary vaccination schedule. |
| **Covariates** | |
| Ethnicity | *The Civil Registration System*[1]. Defined by registered place of birth and categorized according Nordic (Denmark, Finland, Norway, or Sweden), Western (rest of Europe, united states, Australia or New Zealand), and non-Western countries (all others). |
| Region of residency | *The Civil Registration System*[1]. Defined by the last registered address at baseline (1 January 2021) and grouped according: Northern Denmark Region, Central Denmark Region, Region of Southern Denmark, Capital Region of Denmark, and Region Zealand. |
| Vaccination priority groups | *The Danish Vaccination Register*[2]. The Danish Health Agency have provided the governmentally assigned Covid-19 vaccine priority groups that were among other prioritized according to high risk of severe Covid-19 (yes/no; last update 24 May 2021). |
| **Comorbidities** | *The National Patient Register*[3]. Defined as primary diagnoses assigned by the treating physician and registered according to ICD-10 classification system regardless of type of hospital contact registered between 1 January 2018 and baseline (1 January 2021) and included as a categorized sum (0, 1, or ≥2 comorbidities). |
| Asthma | ICD-10 codes: J45, J46 |
| Chronic respiratory disorder | ICD-10 codes: E84, J41-J44, J47, J84 |
| Chronic cardiac disorder | ICD-10 codes: I05-I08, I20-I28, I34-I37, I42-I51 |
| Renal disorder | ICD-10 codes: N03, N05, N07, N18, N19, N25, N26, N27 |
| Diabetes | ICD-10 codes: E10-E14 |
| Autoimmune disorder | ICD-10 codes: D510, D590, D591, D690, D693, D86, E035, E039, E050, E055, E059, E063, E065, E271, E272, E310, G04, G131, G35, G36, G61, G700, H20, I00, I02, K50, |

| | |
|---|---|
| | K51, K732, K743, K900, L10, L12, L130, L40, L63, L80, M05, M06, M08, M30, M311, M313, M315, M316, M317, M32-M34, M350, M351-M353, M358, M359, M45, M60 |
| Epilepsy | ICD-10 codes: G40, G41 |
| Malignancy | ICD-10 codes: C00-C96 (not C44), D70-D72, D730, D81-D84 |
| Psychiatric disorder | ICD-10 codes: F00-F99 |

ICD-10 denotes International Classification of Diseases System, version 10. [a]Similarly, we censored individuals vaccinated with a fourth dose prior to 15 September 2022 (first day of national rollout of the fourth vaccine dose to the general population). [b]Similarly, we censored individuals receiving a fourth dose within 90 days of the third dose. We also excluded individuals with history of the respective outcome under studied prior to the follow-up period.

**Supplementary Table S2. Outcome definitions.**

| Outcomes | ICD-10 codes |
|---|---|
| Anaphylaxis | T782, T783, T805, T886 |
| Ischemic cardiac event | I20-I251 |
| Cerebrovascular event | I60-66, G450-G453 |
| Cerebrovascular infarction (incl. TIA)[b] | I63, I64, G450-453 |
| Arterial thromboembolism | I74 |
| Deep venous thrombosis | I80-82 (not I800, I808C, or I821) |
| Pulmonary embolism | I26 |
| Myocarditis or pericarditis | I300, I308, I309, I328, I401, I408, I409, I418, I514 |
| Myocarditis[b] | I401, I408, I409, I418, I514 |
| Pericarditis[b] | I300, I308, I309, I328 |
| Cerebral venous thrombosis | I636, I676 |
| Thrombocytopenia or coagulative disorders | D65, D683, D686, D688-689, D690, D693-D699 (not D697 or D698A) |
| Guillain-Barré syndrome | G610 |
| Bell's palsy | G510 |
| Transverse myelitis | G373 |
| Encephalomyelitis or encephalitis | G040, G040A, G048, G049, G058, G361 |
| Narcolepsy | G474 |
| Appendicitis | K35-K37 |
| Aseptic arthritis | M10, M119, M130, M131, M139 |
| Type 1 diabetes mellitus | E10 |
| Subacute thyroiditis | E061 |
| Heart failure | I110, I420, I426-I429, I50, J81, |
| Arrythmia | I44-I49 |
| Acute liver failure | K71, K72 |
| Acute kidney failure | D593, I12, I13, N00-N02, N04-N05, N08, N10, N141, N142, N144, N17, N19, R34 |
| Acute pancreatitis | K850, K853, K858, K859 |
| Erythema multiforme | L51 |
| Seizure | G40, G41 |
| Arterial Aneurysm | I71, I72 |
| Uveitis | H20, H30 |

ICD-10 denotes International Classification of Diseases System, version 10, TIA transient cerebral ischemic attack, and NA not applicable; information on death was gathered from the Danish Civil Registration System. [a]Diagnoses were identified through use of the Danish National Patient Register. Only diagnoses registered as primary diagnoses were included in the outcome definitions, however, secondary diagnoses were included in a sensitivity analysis. Main outcome definition included any hospital contact, whereas in a sensitivity analysis we defined severe adverse events as by hospital contacts with a duration of ≥5 hours [b]Examined separately post-hoc.

**Supplementary Figure S1. Schematic figure of the study design.**

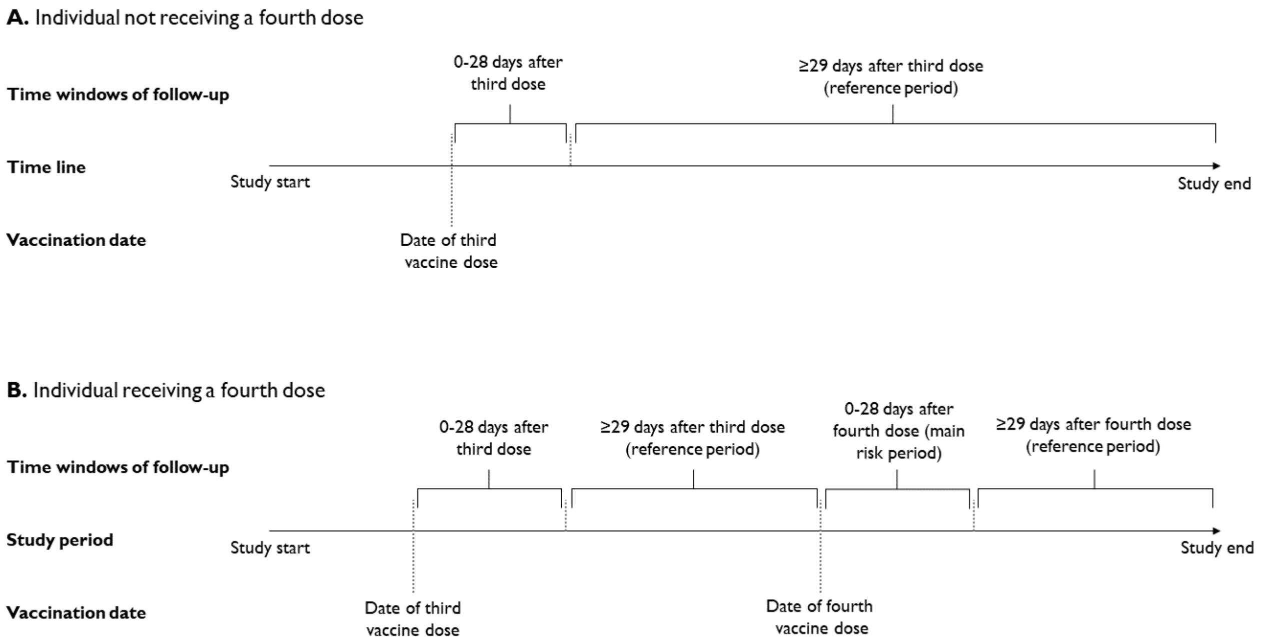

**Supplementary Table S3: Number of events and person-years within 28 days after vaccination with a bivalent omicron-containing vaccine as a fourth dose by type of vaccine and reference period in Danish 50+ year-olds during 15 September 2022 to 10 December 2022.**[a]

| Outcome event | Number of events / person-years | | | |
|---|---|---|---|---|
| | BNT BA.4-5-containing | BNT BA.1-containing | MOD BA.1-containing | Reference period |
| Anaphylaxis | 22 / 85780.6 | 7 / 40100.1 | 1 / 4278.8 | 363 / 1891644.8 |
| Ischemic cardiac event | 425 / 83391.3 | 227 / 38670.0 | 20 / 4134.8 | 9992 / 1838445.5 |
| Cerebrovascular event | 468 / 84037.0 | 305 / 38670.9 | 25 / 4176.9 | 11523 / 1847731.6 |
| Arterial thromboembolism | 17 / 85808.8 | 4 / 40112.2 | 0 / 4280.3 | 385 / 1892133.7 |
| Deep vein thrombosis | 140 / 85065.6 | 90 / 39674.8 | 7 / 4247.2 | 3593 / 1875648.1 |
| Pulmonary embolism | 125 / 85349.3 | 86 / 39774.9 | 5 / 4253.2 | 3336 / 1881117.3 |
| Myocarditis or pericarditis | 18 / 85799.8 | 9 / 40116.3 | 2 / 4280.8 | 322 / 1892114.4 |
| Cerebral venous thrombosis | 2 / 85864.3 | 1 / 40148.7 | 0 / 4283.3 | 37 / 1893510.4 |
| Thrombocytopenia and coagulative disorders | 18 / 85748.0 | 10 / 40077.3 | 2 / 4277.4 | 489 / 1890679.3 |
| Guillain-Barré syndrome | 1 / 85860.5 | 0 / 40147.6 | 0 / 4283.4 | 48 / 1893425.3 |
| Bell's palsy | 19 / 85777.8 | 9 / 40106.3 | 1 / 4278.7 | 449 / 1891679.3 |
| Transverse myelitis | NE | NE | NE | 13 / 1893596.9 |
| Encephalomyelitis or encephalitis | 6 / 85857.0 | 2 / 40141.9 | 1 / 4283.1 | 67 / 1893277.3 |
| Narcolepsy | NE | NE | NE | 7 / 1893483.6 |
| Appendicitis | 81 / 85457.2 | 39 / 39953.9 | 6 / 4263.6 | 1911 / 1885284.3 |
| Aseptic arthritis | 123 / 85007.9 | 87 / 39608.3 | 5 / 4238.1 | 3213 / 1873648.6 |
| Type 1 diabetes mellitus | 13 / 85188.1 | 10 / 39809.1 | 1 / 4244.2 | 559 / 1878618.9 |
| Subacute thyroiditis | 10 / 85841.3 | 6 / 40141.7 | 0 / 4282.4 | 138 / 1893045.7 |
| Heart failure | 305 / 84375.4 | 200 / 39005.9 | 8 / 4196.3 | 8409 / 1855723.5 |
| Arrythmia | 925 / 80698.5 | 561 / 36316.3 | 44 / 3943.1 | 21642 / 1769323.7 |
| Acute liver failure | 7 / 85821.6 | 3 / 40122.7 | 0 / 4280.1 | 375 / 1892421.8 |
| Acute kidney failure | 124 / 85300.9 | 103 / 39704.3 | 9 / 4245.7 | 3819 / 1878542.3 |
| Acute pancreatitis | 43 / 85654.8 | 30 / 40021.5 | 4 / 4272.8 | 1256 / 1888827.6 |

**Supplementary Table S3: Number of events and person-years within 28 days after vaccination with a bivalent omicron-containing vaccine as a fourth dose by type of vaccine and reference period in Danish 50+ year-olds during 15 September 2022 to 10 December 2022.[a]**

| | Number of events / person-years | | | |
|---|---|---|---|---|
| **Outcome event** | **BNT BA.4-5-containing** | **BNT BA.1-containing** | **MOD BA.1-containing** | **Reference period** |
| Erythema multiforme | 1 / 85866.0 | 0 / 40148.9 | 0 / 4283.6 | 36 / 1893528.2 |
| Seizure | 55 / 85326.4 | 53 / 39735.5 | 3 / 4254.2 | 1788 / 1879652.3 |
| Arterial aneurysm | 180 / 85021.6 | 106 / 39579.0 | 12 / 4228.2 | 3566 / 1874302.8 |
| Uveitis | 19 / 85722.3 | 12 / 40077.7 | 0 / 4277.7 | 622 / 1890418.3 |

BNT denotes BNT162b2, MOD mRNA-1273, and NE not estimable. [a]The corresponding incidence rate ratios with 95% confidence intervals are shown in Figure 2.

**Supplementary Table S4: Risk of adverse events within 28 days after vaccination with a bivalent omicron-containing mRNA-booster vaccine as the fourth dose in Danish 50+ year-olds during 15 September 2022 to 10 December 2022 using an alternative analytical approach of a self-controlled case series design.**

| | Number of events / person-years | | |
|---|---|---|---|
| **Outcome event** | **Bivalent boosters** | **Reference period** | **Incidence rate ratio (95% CI)** |
| Anaphylaxis | 30 / 47.6 | 347 / 673.7 | 1.05 (0.67-1.65) |
| Ischemic cardiac event | 672 / 1354.5 | 9726 / 18814.4 | 0.92 (0.84-1.01) |
| Cerebrovascular event | 798 / 1329.4 | 11291 / 19290.0 | 0.96 (0.89-1.05) |
| Arterial thromboembolism | 21 / 38.8 | 371 / 621.2 | 1.15 (0.67-1.96) |
| Deep vein thrombosis | 237 / 448.4 | 3460 / 6502.5 | 1.01 (0.86-1.18) |
| Pulmonary embolism | 216 / 369.8 | 3234 / 5518.4 | 0.85 (0.72-1.00) |
| Myocarditis or pericarditis | 29 / 38.6 | 314 / 597.8 | 1.52 (0.94-2.45) |
| Cerebral venous thrombosis | 3 / 4.5 | 36 / 64.3 | 1.49 (0.31-7.10) |
| Thrombocytopenia or coagulative disorder | 30 / 52.9 | 466 / 834.7 | 0.87 (0.57-1.34) |
| Guillain-Barré syndrome | 1 / 4.4 | 48 / 71.5 | 0.20 (0.03-1.57) |
| Bell's palsy | 29 / 53.9 | 435 / 793.4 | 0.96 (0.62-1.51) |
| Transverse myelitis | 0 / 1.9 | 13 / 26.7 | NE |
| Encephalomyelitis or encephalitis | 9 / 7.7 | 66 / 125.1 | 1.66 (0.68-4.07) |
| Narcolepsy | 0 / 1.0 | 6 / 14.3 | NE |
| Appendicitis | 126 / 260.6 | 1856 / 3607.4 | 0.97 (0.78-1.21) |
| Aseptic arthritis | 215 / 431.6 | 3090 / 6130.2 | 0.92 (0.78-1.09) |
| Type 1 diabetes mellitus | 24 / 72.0 | 538 / 1038.5 | 0.81 (0.50-1.31) |
| Subacute thyroiditis | 16 / 16.6 | 131 / 257.1 | 1.97 (0.99-3.91) |
| Heart failure | 513 / 921.7 | 8160 / 13543.2 | 0.80 (0.72-0.89) |
| Arrythmia | 1530 / 2777.5 | 20846 / 38150.4 | 0.93 (0.87-0.99) |
| Acute liver failure | 10 / 31.6 | 368 / 533.3 | 0.37 (0.19-0.74) |
| Acute kidney failure | 236 / 362.7 | 3736 / 5670.7 | 0.79 (0.68-0.92) |
| Acute pancreatitis | 77 / 146.5 | 1224 / 2107.8 | 0.82 (0.63-1.07) |
| Erythema multiforme | 1 / 3.7 | 36 / 59.7 | 0.43 (0.05-3.76) |
| Seizure | 111 / 210.1 | 1707 / 3053.6 | 0.89 (0.71-1.12) |
| Arterial aneurysm | 298 / 468.9 | 3429 / 6322.4 | 0.94 (0.82-1.09) |
| Uveitis | 31 / 76.8 | 600 / 1124.9 | 0.97 (0.63-1.49) |

NE denotes not estimable.

**Supplementary Table S5: Risk of adverse events within 28 days after vaccination with a bivalent omicron-containing mRNA-booster vaccine as the fourth dose in Danish 50+ year-olds during 15 September 2022 to 10 December 2022 using an alternative analytical approach of an observed vs expected design.**

| | Number of observed / expected events | |
|---|---|---|
| **Outcome event** | **Bivalent boosters** | **Rate ratio (95% CI)** |
| Anaphylaxis | 30 / 38.3 | 0.78 (0.53-1.12) |
| Ischemic cardiac event | 672 / 1123.5 | 0.60 (0.55-0.65) |
| Cerebrovascular event | 798 / 845.2 | 0.94 (0.88-1.01) |
| Arterial thromboembolism | 21 / 36.6 | 0.57 (0.35-0.88) |
| Deep vein thrombosis | 237 / 322.3 | 0.74 (0.64-0.84) |
| Pulmonary embolism | 216 / 269.7 | 0.80 (0.70-0.92) |
| Myocarditis or pericarditis | 29 / 21.6 | 1.35 (0.90-1.93) |
| Cerebral venous thrombosis | 3 / 2.7 | 1.12 (0.23-3.28) |
| Thrombocytopenia or coagulative disorder | 30 / 74.4 | 0.40 (0.27-0.58) |
| Guillain-Barré syndrome | 1 / 5.8 | 0.17 (0.00-0.97) |
| Bell's palsy | 29 / 33.2 | 0.87 (0.59-1.26) |
| Transverse myelitis | 0 / 1.3 | NE |
| Encephalomyelitis or encephalitis | 9 / 7.7 | 1.17 (0.54-2.23) |
| Narcolepsy | 0 / 4.0 | NE |
| Appendicitis | 126 / 110.1 | 1.14 (0.95-1.36) |
| Aseptic arthritis | 215 / 347.5 | 0.62 (0.54-0.71) |
| Type 1 diabetes mellitus | 24 / 402.3 | 0.06 (0.04-0.09) |
| Subacute thyroiditis | 16 / 7.2 | 2.22 (1.27-3.60) |
| Heart failure | 513 / 937.4 | 0.55 (0.50-0.60) |
| Arrythmia | 1530 / 2467.1 | 0.62 (0.59-0.65) |
| Acute liver failure | 10 / 44.4 | 0.23 (0.11-0.41) |
| Acute kidney failure | 236 / 260.0 | 0.91 (0.80-1.03) |
| Acute pancreatitis | 77 / 87.4 | 0.88 (0.70-1.10) |
| Erythema multiforme | 1 / 3.0 | 0.33 (0.01-1.83) |
| Seizure | 111 / 349.3 | 0.32 (0.26-0.38) |
| Arterial aneurysm | 298 / 392.9 | 0.76 (0.67-0.85) |
| Uveitis | 31 / 56.7 | 0.55 (0.37-0.78) |

NE denotes not estimable.

**Supplementary Table S6: Risk of severe adverse events (hospitalization for ≥5 hours) within 28 days after vaccination with a bivalent omicron-containing vaccine as a fourth dose in Danish 50+ year-olds during 15 September 2022 to 10 December 2022.**

| | Number of events / person-years | | |
|---|---|---|---|
| **Outcome event** | **Bivalent boosters** | **Reference period** | **Incidence rate ratio (95% CI)** |
| Anaphylaxis | 20 / 130178 | 217 / 1891849 | 1.16 (0.68-2.01) |
| Ischemic cardiac event | 513 / 126464 | 8188 / 1841576 | 0.90 (0.81-1.00) |
| Cerebrovascular event | 683 / 127072 | 9990 / 1849868 | 0.94 (0.86-1.03) |
| Arterial thromboembolism | 14 / 130212 | 271 / 1892266 | 0.91 (0.49-1.70) |
| Deep vein thrombosis | 88 / 129263 | 1327 / 1878842 | 0.97 (0.76-1.25) |
| Pulmonary embolism | 148 / 129461 | 2556 / 1882073 | 0.75 (0.62-0.91) |
| Myocarditis or pericarditis | 20 / 130204 | 263 / 1892203 | 1.18 (0.69-2.03) |
| Cerebral venous thrombosis | 3 / 130297 | 29 / 1893523 | 2.10 (0.43-10.15) |
| Thrombocytopenia and coagulative disorders | 9 / 130135 | 220 / 1891056 | 0.70 (0.33-1.48) |
| Guillain-Barré syndrome | 1 / 130292 | 41 / 1893436 | 0.23 (0.03-1.84) |
| Bell's palsy | 11 / 130193 | 166 / 1892014 | 1.16 (0.56-2.41) |
| Transverse myelitis | 0 / 130305 | 5 / 1893609 | NE |
| Encephalomyelitis or encephalitis[a] | 9 / 130284 | 50 / 1893304 | 2.67 (1.01-7.07) |
| Appendicitis | 126 / 129677 | 1897 / 1885316 | 1.03 (0.83-1.27) |
| Aseptic arthritis | 40 / 129171 | 786 / 1877282 | 0.78 (0.54-1.12) |
| Type 1 diabetes mellitus | 4 / 129297 | 133 / 1879275 | 0.70 (0.23-2.16) |
| Subacute thyroiditis | 0 / 130280 | 6 / 1893231 | NE |
| Heart failure | 317 / 127945 | 5418 / 1859868 | 0.75 (0.66-0.85) |
| Arrythmia | 904 / 122126 | 12768 / 1782238 | 0.90 (0.83-0.98) |
| Acute liver failure | 9 / 130238 | 260 / 1892577 | 0.45 (0.22-0.93) |
| Acute kidney failure | 188 / 129322 | 3183 / 1879340 | 0.75 (0.64-0.89) |
| Acute pancreatitis | 74 / 129955 | 1209 / 1888900 | 0.82 (0.62-1.07) |
| Erythema multiforme | 0 / 130300 | 19 / 1893553 | NE |
| Seizure | 59 / 129434 | 830 / 1881012 | 0.91 (0.67-1.24) |
| Arterial aneurysm | 76 / 129152 | 1017 / 1877798 | 1.00 (0.76-1.31) |
| Uveitis | 6 / 130142 | 67 / 1891200 | 1.35 (0.49-3.74) |

NE denotes not estimable. [a]The rate ratio for encephalomyelitis or encephalitis was 1.55 (95% CI 0.71-2.95) and 2.77 (95% CI 0.99-7.75) with use of observed vs. expected and self-controlled case series analysis, respectively.

**Supplementary Table S7: Sensitivity analysis of the associated risk of adverse events within 28 days following fourth dose bivalent omicron-containing booster vaccination where including both primary and secondary diagnoses in the outcome definitions.**

| | Number of events / person-years | | |
|---|---|---|---|
| **Outcome** | **Bivalent boosters** | **Reference period** | **Incidence rate ratio (95% CI)** |
| Anaphylaxis | 31 / 130156 | 400 / 1891605 | 0.92 (0.60-1.39) |
| Ischemic cardiac event | 742 / 126093 | 10864 / 1837213 | 0.96 (0.88-1.04) |
| Cerebrovascular event | 875 / 126740 | 12807 / 1846000 | 0.95 (0.88-1.03) |
| Arterial thromboembolism | 22 / 130197 | 425 / 1892073 | 0.88 (0.54-1.45) |
| Deep vein thrombosis | 258 / 128938 | 4069 / 1875052 | 0.93 (0.80-1.08) |
| Pulmonary embolism | 238 / 129339 | 3839 / 1880619 | 0.80 (0.69-0.93) |
| Myocarditis or pericarditis | 32 / 130192 | 379 / 1892054 | 1.28 (0.83-1.97) |
| Cerebral venous thrombosis | 3 / 130296 | 40 / 1893506 | 1.58 (0.36-6.92) |
| Thrombocytopenia and coagulative disorders | 45 / 130087 | 700 / 1890477 | 0.81 (0.57-1.14) |
| Guillain-Barré syndrome | 3 / 130291 | 55 / 1893417 | 0.64 (0.18-2.30) |
| Bell's palsy | 29 / 130160 | 472 / 1891644 | 0.94 (0.61-1.45) |
| Transverse myelitis | 1 / 130303 | 14 / 1893595 | 1.10 (0.10-12.18) |
| Encephalomyelitis or encephalitis | 11 / 130280 | 77 / 1893261 | 1.80 (0.81-4.00) |
| Narcolepsy | 2 / 130295 | 14 / 1893476 | 1.05 (0.19-5.73) |
| Appendicitis | 129 / 129670 | 1953 / 1885224 | 1.01 (0.82-1.25) |
| Aseptic arthritis | 251 / 128767 | 3953 / 1872615 | 0.85 (0.73-0.98) |
| Type 1 diabetes mellitus | 41 / 129197 | 936 / 1878074 | 0.80 (0.56-1.14) |
| Subacute thyroiditis | 17 / 130265 | 143 / 1893038 | 1.60 (0.86-2.98) |
| Heart failure | 609 / 127366 | 10584 / 1853124 | 0.74 (0.68-0.82) |
| Arrythmia | 1903 / 120307 | 27536 / 1761569 | 0.91 (0.87-0.96) |
| Acute liver failure | 15 / 130219 | 498 / 1892336 | 0.43 (0.25-0.75) |
| Acute kidney failure | 386 / 129071 | 6185 / 1876334 | 0.81 (0.72-0.91) |
| Acute pancreatitis | 83 / 129939 | 1358 / 1888695 | 0.80 (0.62-1.03) |
| Erythema multiforme | 1 / 130298 | 41 / 1893522 | 0.40 (0.05-3.36) |
| Seizure | 128 / 129282 | 2084 / 1879244 | 0.87 (0.71-1.07) |
| Arterial aneurysm | 335 / 128752 | 4149 / 1873400 | 0.95 (0.83-1.08) |
| Uveitis | 32 / 130073 | 653 / 1890363 | 0.85 (0.57-1.28) |

**Supplementary Table S8. Risk of adverse events after vaccination with a bivalent omicron-containing mRNA-booster vaccine as the fourth dose in Danish 50+ year-olds during 15 September 2022 to 10 December 2022 with use of alternative risk periods.**

| Alternative risk period/outcome | Number of events / person-years | | Incidence rate ratio (95% CI) |
|---|---|---|---|
| | Bivalent boosters | Reference period | |
| **Risk period restricted to the first 7 days after fourth vaccine dose** | | | |
| Anaphylaxis | 7 / 33083 | 398 / 2115427 | 0.93 (0.43-2.00) |
| Ischemic cardiac event | 148 / 32081 | 11217 / 2055979 | 0.83 (0.70-0.98) |
| Cerebrovascular event | 166 / 32256 | 12902 / 2066397 | 0.79 (0.67-0.92) |
| Arterial thromboembolism | 6 / 33093 | 428 / 2115978 | 1.09 (0.47-2.50) |
| Deep vein thrombosis | 60 / 32786 | 3998 / 2097555 | 0.97 (0.74-1.26) |
| Pulmonary embolism | 35 / 32886 | 3768 / 2103658 | 0.55 (0.39-0.77) |
| Myocarditis or pericarditis | 9 / 33092 | 368 / 2115955 | 1.55 (0.77-3.09) |
| Cerebral venous thrombosis | 2 / 33117 | 43 / 2117512 | 4.43 (0.89-22.12) |
| Thrombocytopenia or coagulative disorder | 9 / 33068 | 536 / 2114346 | 0.96 (0.49-1.89) |
| Guillain-Barré syndrome | 0 / 33116 | 56 / 2117417 | NE |
| Bell's palsy | 10 / 33084 | 495 / 2115469 | 1.36 (0.71-2.62) |
| Transverse myelitis | 0 / 33119 | 13 / 2117610 | NE |
| Encephalomyelitis or encephalitis | 1 / 33114 | 79 / 2117253 | 0.56 (0.07-4.12) |
| Narcolepsy | 0 / 33117 | 7 / 2117482 | NE |
| Appendicitis | 33 / 32960 | 2118 / 2108328 | 1.04 (0.73-1.48) |
| Aseptic arthritis | 56 / 32752 | 3616 / 2095306 | 0.94 (0.71-1.23) |
| Type 1 diabetes mellitus | 12 / 32850 | 611 / 2100857 | 1.72 (0.94-3.15) |
| Subacute thyroiditis | 4 / 33109 | 162 / 2116997 | 1.41 (0.50-3.96) |
| Heart failure | 127 / 32430 | 9345 / 2075325 | 0.82 (0.69-0.98) |
| Arrythmia | 352 / 30760 | 24219 / 1978552 | 0.86 (0.77-0.95) |
| Acute liver failure | 4 / 33099 | 404 / 2116300 | 0.73 (0.27-2.01) |
| Acute kidney failure | 45 / 32853 | 4213 / 2100785 | 0.61 (0.45-0.83) |
| Acute pancreatitis | 19 / 33029 | 1392 / 2112285 | 0.84 (0.52-1.33) |
| Erythema multiforme | 0 / 33118 | 42 / 2117532 | NE |
| Seizure | 34 / 32869 | 1985 / 2102039 | 1.08 (0.76-1.54) |
| Arterial aneurysm | 62 / 32748 | 4025 / 2095996 | 0.81 (0.62-1.04) |
| Uveitis | 4 / 33062 | 689 / 2114067 | 0.45 (0.16-1.21) |
| **Risk period restricted to the first 14 days after fourth vaccine dose** | | | |
| Anaphylaxis | 14 / 65919 | 388 / 2040463 | 0.92 (0.52-1.62) |
| Ischemic cardiac event | 319 / 63920 | 10833 / 1983109 | 0.89 (0.79-1.00) |

**Supplementary Table S8. Risk of adverse events after vaccination with a bivalent omicron-containing mRNA-booster vaccine as the fourth dose in Danish 50+ year-olds during 15 September 2022 to 10 December 2022 with use of alternative risk periods.**

| Alternative risk period/outcome | Number of events / person-years | | Incidence rate ratio (95% CI) |
|---|---|---|---|
| | Bivalent boosters | Reference period | |
| Cerebrovascular event | 386 / 64268 | 12436 / 1993146 | 0.92 (0.83-1.03) |
| Arterial thromboembolism | 12 / 65940 | 415 / 2040993 | 1.11 (0.60-2.06) |
| Deep vein thrombosis | 120 / 65328 | 3863 / 2023219 | 0.97 (0.80-1.17) |
| Pulmonary embolism | 103 / 65526 | 3614 / 2029108 | 0.82 (0.66-1.00) |
| Myocarditis or pericarditis | 20 / 65938 | 351 / 2040972 | 1.88 (1.13-3.11) |
| Cerebral venous thrombosis | 3 / 65988 | 38 / 2042474 | 3.78 (0.88-16.17) |
| Thrombocytopenia or coagulative disorder | 18 / 65891 | 516 / 2039421 | 0.96 (0.58-1.58) |
| Guillain-Barré syndrome | 0 / 65986 | 55 / 2042383 | NE |
| Bell's palsy | 15 / 65921 | 480 / 2040502 | 1.00 (0.58-1.73) |
| Transverse myelitis | 0 / 65992 | 13 / 2042568 | NE |
| Encephalomyelitis or encephalitis | 4 / 65981 | 76 / 2042224 | 1.18 (0.40-3.47) |
| Narcolepsy | 0 / 65988 | 7 / 2042446 | NE |
| Appendicitis | 68 / 65675 | 2044 / 2033610 | 1.08 (0.84-1.40) |
| Aseptic arthritis | 109 / 65260 | 3478 / 2021054 | 0.90 (0.74-1.11) |
| Type 1 diabetes mellitus | 17 / 65455 | 594 / 2026411 | 1.20 (0.71-2.02) |
| Subacute thyroiditis | 8 / 65972 | 153 / 2041976 | 1.46 (0.67-3.17) |
| Heart failure | 259 / 64616 | 9035 / 2001763 | 0.83 (0.72-0.94) |
| Arrythmia | 744 / 61282 | 23357 / 1908464 | 0.90 (0.83-0.97) |
| Acute liver failure | 8 / 65952 | 389 / 2041304 | 0.72 (0.35-1.51) |
| Acute kidney failure | 115 / 65460 | 4079 / 2026337 | 0.78 (0.64-0.95) |
| Acute pancreatitis | 37 / 65813 | 1348 / 2037430 | 0.80 (0.57-1.13) |
| Erythema multiforme | 0 / 65989 | 41 / 2042494 | NE |
| Seizure | 53 / 65492 | 1931 / 2027542 | 0.82 (0.61-1.09) |
| Arterial aneurysm | 147 / 65250 | 3862 / 2021732 | 0.97 (0.81-1.15) |
| Uveitis | 9 / 65877 | 672 / 2039147 | 0.49 (0.25-0.96) |
| **Risk period extended to 90 days after fourth vaccine dose** | | | |
| Anaphylaxis | 57 / 262345 | 272 / 1384066 | 0.85 (0.56-1.29) |
| Ischemic cardiac event | 1381 / 254126 | 7156 / 1344999 | 0.92 (0.84-1.00) |
| Cerebrovascular event | 1623 / 255346 | 8439 / 1351895 | 0.85 (0.78-0.92) |
| Arterial thromboembolism | 34 / 262429 | 291 / 1384423 | 0.49 (0.30-0.81) |

**Supplementary Table S8. Risk of adverse events after vaccination with a bivalent omicron-containing mRNA-booster vaccine as the fourth dose in Danish 50+ year-olds during 15 September 2022 to 10 December 2022 with use of alternative risk periods.**

| Alternative risk period/outcome | Number of events / person-years | | Incidence rate ratio (95% CI) |
|---|---|---|---|
| | Bivalent boosters | Reference period | |
| Deep vein thrombosis | 471 / 259910 | 2633 / 1372343 | 0.85 (0.74-0.99) |
| Pulmonary embolism | 462 / 260687 | 2337 / 1376399 | 0.76 (0.65-0.88) |
| Myocarditis or pericarditis | 47 / 262424 | 240 / 1384406 | 0.94 (0.59-1.50) |
| Cerebral venous thrombosis | 4 / 262627 | 28 / 1385436 | 0.79 (0.18-3.56) |
| Thrombocytopenia or coagulative disorder | 65 / 262228 | 364 / 1383377 | 0.66 (0.45-0.96) |
| Guillain-Barré syndrome | 4 / 262617 | 41 / 1385374 | 0.26 (0.08-0.86) |
| Bell's palsy | 58 / 262356 | 358 / 1384079 | 0.92 (0.61-1.40) |
| Transverse myelitis | 0 / 262642 | 8 / 1385498 | NE |
| Encephalomyelitis or encephalitis | 10 / 262596 | 52 / 1385265 | 0.48 (0.21-1.12) |
| Narcolepsy | 0 / 262626 | 7 / 1385415 | NE |
| Appendicitis | 239 / 261365 | 1415 / 1379352 | 0.87 (0.71-1.06) |
| Aseptic arthritis | 436 / 259638 | 2409 / 1370916 | 0.74 (0.64-0.86) |
| Type 1 diabetes mellitus | 51 / 260485 | 404 / 1374537 | 0.73 (0.48-1.10) |
| Subacute thyroiditis | 21 / 262567 | 106 / 1385086 | 0.77 (0.41-1.47) |
| Heart failure | 1189 / 256874 | 6121 / 1357746 | 0.79 (0.72-0.88) |
| Arrythmia | 3246 / 242808 | 15767 / 1294964 | 0.91 (0.85-0.96) |
| Acute liver failure | 29 / 262479 | 262 / 1384634 | 0.42 (0.26-0.69) |
| Acute kidney failure | 507 / 260415 | 2861 / 1374549 | 0.64 (0.56-0.74) |
| Acute pancreatitis | 159 / 261907 | 916 / 1381989 | 0.70 (0.55-0.89) |
| Erythema multiforme | 4 / 262630 | 27 / 1385450 | 0.83 (0.16-4.32) |
| Seizure | 255 / 260574 | 1278 / 1375293 | 0.99 (0.80-1.22) |
| Arterial aneurysm | 629 / 259535 | 2547 / 1371468 | 1.03 (0.90-1.19) |
| Uveitis | 68 / 262182 | 466 / 1383147 | 1.02 (0.69-1.52) |

NE denotes not estimable.

**Supplementary Table S9:** Risk of cerebrovascular infarction, **myocarditis and/or pericarditis after vaccination with the individual bivalent omicron-containing vaccine types as a fourth dose by sex and age groups within 28 days in Danish 50+ year-olds during 15 September 2022 to 10 December 2022.**

| Outcome/subgroups | BNT BA.4-5-containing | | BNT BA.1-containing | | MOD BA.1-containing | | Reference period | | Incidence rate ratio (95% CI) | | |
|---|---|---|---|---|---|---|---|---|---|---|---|
| | Events/ PYRS | Rate/ 1,000,000 | Events / PYRS | Rate/ 1,000,000 | Events / PYRS | Rate/ 1,000,000 | Events/ PYRS | Rate/ 1,000,000 | BNT BA.4-5-containing | BNT BA.1-containing | MOD BA.1-containing |
| **Cerebrovascular infarction (including TIA)** | | | | | | | | | | | |
| All | 377 / 84259 | 343.0 | 246 / 38856 | 485.3 | 21 / 4189 | 384.3 | 9687 / 1853451 | 400.7 | 0.97 (0.86-1.09) | 0.98 (0.85-1.12) | 0.83 (0.54-1.29) |
| Females | 156 / 43643 | 274.0 | 118 / 21860 | 413.8 | 16 / 2292 | 535.2 | 4317 / 974392 | 339.6 | 0.92 (0.77-1.10) | 0.91 (0.74-1.11) | 1.27 (0.76-2.13) |
| Males | 221 / 40616 | 417.1 | 128 / 16997 | 577.3 | 5 / 1897 | 202.0 | 5370 / 879059 | 468.3 | 1.01 (0.87-1.19) | 1.04 (0.86-1.26) | 0.41 (0.17-0.99) |
| Aged 50 to 64 years | 75 / 39699 | 144.8 | 21 / 8206 | 196.2 | 2 / 1315 | 116.6 | 1716 / 806017 | 163.2 | 0.89 (0.69-1.15) | 1.20 (0.77-1.87) | 0.75 (0.19-3.03) |
| Aged 65 to 79 years | 180 / 32245 | 427.9 | 110 / 18751 | 449.7 | 6 / 1464 | 314.1 | 4036 / 690056 | 448.4 | 0.95 (0.81-1.13) | 1.02 (0.83-1.25) | 0.74 (0.33-1.65) |
| Aged 80 years or older | 99 / 7984 | 950.6 | 105 / 9726 | 827.6 | 12 / 1192 | 771.5 | 3351 / 266105 | 965.4 | 1.07 (0.86-1.33) | 0.92 (0.74-1.13) | 0.94 (0.53-1.66) |
| **Myocarditis or pericarditis** | | | | | | | | | | | |
| All[a] | 18 / 85800 | 16.1 | 9 / 40116 | 17.2 | 2 / 4281 | 35.8 | 322 / 1892114 | 13.0 | 1.05 (0.58-1.92) | 1.29 (0.61-2.74) | 3.17 (0.77-13.11) |
| Females | 8 / 44256 | 13.9 | 5 / 22443 | 17.1 | 2 / 2331 | 65.8 | 130 / 990933 | 10.1 | 0.87 (0.33-2.33) | 1.55 (0.58-4.10) | 6.45 (1.50-27.82) |
| Males | 10 / 41544 | 18.5 | 4 / 17673 | 17.4 | 0 / 1950 | NE | 192 / 901182 | 16.3 | 1.20 (0.56-2.58) | 0.99 (0.30-3.28) | NE |
| Aged 50 to 64 years | 8 / 40022 | 15.3 | 2 / 8313 | 18.4 | 1 / 1327 | 57.8 | 144 / 812580 | 13.6 | 1.30 (0.57-2.95) | 1.46 (0.34-6.23) | 5.37 (0.72-40.12) |
| Aged 65 to 79 years | 6 / 32993 | 13.9 | 4 / 19283 | 15.9 | 1 / 1494 | 51.3 | 117 / 706700 | 12.7 | 0.95 (0.38-2.38) | 1.04 (0.35-3.04) | 3.42 (0.46-25.70) |

**Supplementary Table S9:** Risk of cerebrovascular infarction, **myocarditis and/or pericarditis after vaccination with the individual bivalent omicron-containing vaccine types as a fourth dose by sex and age groups within 28 days in Danish 50+ year-olds during 15 September 2022 to 10 December 2022.**

| | BNT BA.4-5-containing | | BNT BA.1-containing | | MOD BA.1-containing | | Reference period | | Incidence rate ratio (95% CI) | | |
|---|---|---|---|---|---|---|---|---|---|---|---|
| **Outcome/subgroups** | **Events/ PYRS** | **Rate/ 1,000,000** | **Events / PYRS** | **Rate/ 1,000,000** | **Events / PYRS** | **Rate/ 1,000,000** | **Events/ PYRS** | **Rate/ 1,000,000** | **BNT BA.4-5-containing** | **BNT BA.1-containing** | **MOD BA.1-containing** |
| Aged 80 years or older | 0 / 8355 | NE | 2 / 10272 | 14.9 | 0 / 1239 | NE | 41 / 279247 | 11.3 | NE | 1.52 (0.31-7.38) | NE |
| **Myocarditis** | | | | | | | | | | | |
| All | 6 / 85859 | 5.4 | 2 / 40146 | 3.8 | 1 / 4283 | 17.9 | 64 / 1893405 | 2.6 | 3.74 (1.16-12.05) | 2.59 (0.52-12.94) | 12.39 (1.46-105.47) |
| Females | 3 / 44278 | 5.2 | 2 / 22455 | 6.8 | 1 / 2332 | 32.9 | 29 / 991433 | 2.2 | 7.10 (1.06-47.81) | 8.50 (1.24-58.44) | 59.58 (4.97-714.53) |
| Males | 3 / 41581 | 5.5 | 0 / 17690 | NE | 0 / 1951 | NE | 35 / 901972 | 3.0 | 2.42 (0.54-10.83) | NE | NE |
| Aged 50 to 64 years | 3 / 40050 | 5.7 | 0 / 8320 | NE | 1 / 1327 | 57.8 | 25 / 813144 | 2.4 | 4.76 (0.80-28.37) | NE | 50.46 (4.55-559.52) |
| Aged 65 to 79 years | 3 / 33017 | 7.0 | 1 / 19297 | 4.0 | 0 / 1495 | NE | 26 / 707193 | 2.8 | 3.04 (0.62-15.00) | 1.55 (0.16-14.66) | NE |
| Aged 80 years or older | 0 / 8361 | NE | 1 / 10279 | 7.5 | 0 / 1240 | NE | 7 / 279421 | 1.9 | NE | 16.95 (0.62-459.83) | NE |
| **Pericarditis** | | | | | | | | | | | |
| All | 14 / 85811 | 12.5 | 7 / 40123 | 13.4 | 1 / 4281 | 17.9 | 265 / 1892351 | 10.7 | 0.85 (0.43-1.71) | 1.12 (0.48-2.64) | 1.82 (0.25-13.31) |
| Females | 7 / 44260 | 12.1 | 3 / 22445 | 10.2 | 1 / 2331 | 32.9 | 104 / 991018 | 8.0 | 0.75 (0.25-2.21) | 0.99 (0.29-3.34) | 3.14 (0.42-23.64) |
| Males | 7 / 41551 | 12.9 | 4 / 17677 | 17.3 | 0 / 1951 | NE | 161 / 901332 | 13.7 | 0.96 (0.39-2.38) | 1.23 (0.37-4.13) | NE |

**Supplementary Table S9: Risk of cerebrovascular infarction, myocarditis and/or pericarditis after vaccination with the individual bivalent omicron-containing vaccine types as a fourth dose by sex and age groups within 28 days in Danish 50+ year-olds during 15 September 2022 to 10 December 2022.**

| | BNT BA.4-5-containing | | BNT BA.1-containing | | MOD BA.1-containing | | Reference period | | Incidence rate ratio (95% CI) | | |
|---|---|---|---|---|---|---|---|---|---|---|---|
| **Outcome/subgroups** | **Events/ PYRS** | **Rate/ 1,000,000** | **Events / PYRS** | **Rate/ 1,000,000** | **Events / PYRS** | **Rate/ 1,000,000** | **Events/ PYRS** | **Rate/ 1,000,000** | **BNT BA.4-5-containing** | **BNT BA.1-containing** | **MOD BA.1-containing** |
| Aged 50 to 64 years | 6 / 40028 | 11.5 | 2 / 8314 | 18.4 | 0 / 1327 | NE | 122 / 812683 | 11.5 | 1.09 (0.43-2.73) | 1.64 (0.38-7.07) | NE |
| Aged 65 to 79 years | 4 / 32998 | 9.3 | 3 / 19286 | 11.9 | 1 / 1494 | 51.3 | 92 / 706793 | 10.0 | 0.75 (0.25-2.25) | 0.94 (0.27-3.20) | 4.09 (0.54-31.00) |
| Aged 80 years or older | 0 / 8356 | NE | 1 / 10274 | 7.5 | 0 / 1239 | NE | 35 / 279276 | 9.6 | NE | 0.78 (0.09-6.47) | NE |

BNT denotes BNT162b2, MOD mRNA-1273, NE not estimable, and PYRS total person-years. Rate/1,000,000 is the incidence rate per 1,000,000 vaccinated within 28 days. [a]The IRR estimates are identical to those reported in Figure 2.
